# Analytical Performance and 99th Percentile Upper Reference Limit of the Novel SPINCHIP High-Sensitivity Cardiac Troponin I Point-of-Care Assay

**DOI:** 10.64898/2026.07.17.26357157

**Authors:** Jenny MacKenzie, Kristin Moberg Aakre, Didrik Paus, Marianne Nordlund Broughton, Gro Leite Storvold, Anniken Olberg, Sebastian Stenmark, Birgitte Boonstra Booij, Sara Scott, Sandrine Michel-Busseret, Lydia Octave, Arnljot Tveit, Magnus N. Lyngbakken, Johan Nilsson, Helge Røsjø

## Abstract

**BACKGROUND:** In line with International Federation of Clinical Chemistry and Laboratory Medicine (IFCC) recommendations for high-sensitivity cardiac troponin assays, analytical validation and reference limit assessments are required to confirm that an assay meets performance criteria. This study evaluated the analytical performance and established the 99^th^ percentile upper reference limit (URL) for the SPINCHIP^®^ High-Sensitivity Cardiac Troponin I (“SPINCHIP hs-cTnI”) point-of-care assay.

**METHODS:** Analytical performance characteristics, including the limit of blank (LoB), limit of detection (LoD), and limit of quantification (LoQ), were assessed. Additionally, 1053 plasma samples and 1055 whole-blood samples were used to determine the URL. Imprecision around the 99^th^ percentile URL was evaluated as part of the analytical validation. High-sensitivity criteria were assessed by confirming measurable cTnI in ≥50% of healthy individuals (n=432 plasma; n=431 whole blood) and achieving imprecision <10% at the 99th percentile (plasma, n=960; whole blood, n=480).

**RESULTS:** SPINCHIP hs-cTnI demonstrated a LoB of 0.3 ng/L; LoDs of 0.8 ng/L (plasma) and 0.9 ng/L (whole blood); and LoQs of 1.1 ng/L (plasma) and 1.4 ng/L (whole blood). The analytical measuring range was 1.1–9000 ng/L. Imprecision at the common 99th percentile URL (14 ng/L) was 5.8%; for men (URL=16 ng/L) 5.6% and for women (URL=10 ng/L) 6.3%. Greater than 85.2% (94.0% and 76.1% in men and women, respectively) of healthy individuals showed measurable cTnI above the LoD.

**CONCLUSIONS:** The SPINCHIP hs-cTnI assay meets the IFCC high-sensitivity requirements, demonstrating <10% imprecision at the 99th percentile, reliable low-concentration precision. and cTnI detection in more than half of healthy individuals.

**IMPACT STATEMENT:** High-sensitivity cardiac troponin assays must demonstrate exceptional analytical rigor to ensure clinicians can rely on them for timely and accurate detection of myocardial injury. This study provides a comprehensive analytical validation of the SPINCHIP^®^ hs-cTnI point-of-care assay to confirm that it meets key International Federation of Clinical Chemistry and Laboratory Medicine high-sensitivity criteria, including precise quantification at the 99th percentile and detection of measurable cardiac troponin concentrations in the majority of healthy individuals. These findings establish the assay’s analytical robustness and support its potential to deliver rapid, trustworthy cardiac troponin results at the point-of-care, helping streamline clinical decision making where speed and accuracy are critical.

## INTRODUCTION

Coronary artery disease (CAD), a major subset of cardiovascular disease, is characterized by atherosclerotic narrowing or obstruction of the coronary arteries and is the most common form of heart disease and a leading cause of global mortality.(1, 2) Acute coronary syndrome (ACS) is the unstable clinical manifestation of CAD, typically presenting with acute chest pain resulting from an abrupt reduction in coronary blood flow caused by plaque rupture, thrombus formation, or other forms of acute coronary obstruction.(2, 3) ACS affects an estimated seven million individuals globally each year.(4)

Approximately 30% of ACS presentations involve ST-elevation myocardial infarction (STEMI), which is diagnosed on the basis of characteristic symptoms, physical examination, and electrocardiographic (ECG) findings.(5) In contrast, patients with non–ST-elevation myocardial infarction (NSTEMI) can be normal or may only demonstrate subtle ECG abnormalities, and definitive diagnosis requires biomarker confirmation based on a rise and/or fall in cardiac troponin concentrations on serial measurements. Cardiac troponin (cTn) is a highly specific biomarker of myocardial injury,(6–9) and high-sensitivity cardiac troponin assays are the recommended method for evaluating suspected acute myocardial infarction (AMI).(9–13) In the absence of cardiac troponin elevation, patients with ischemic symptoms and no persistent ST-segment elevation are classified as having unstable angina.(10, 14, 15)

Because hs-cTn assays differ in analytical design and performance, assay-specific decision thresholds must be established before clinical use. International guidelines recommend use of the 99th percentile upper reference limit (URL), derived from rigorously-defined overall and sex-specific healthy reference populations.(10, 14, 15) The *Fourth Universal Definition of Myocardial Infarction* defines AMI as a rise and/or fall in cTn values, with at least one value exceeding the 99th percentile URL, in the setting of myocardial ischemia.(15) In addition, the 2018 IFCC/AACC consensus statement outlines analytical criteria for hs-cTn assays, including a coefficient of variation (CV) ≤10% at the 99th percentile and measurable cTn concentrations above the limit of detection (LoD) in at least 50% of healthy individuals from a sex-balanced reference cohort.(5, 16–18)

The objective of the present study was to establish overall and sex-specific 99th percentile URLs for the SPINCHIP^®^ High-Sensitivity Cardiac Troponin I point-of-care (SPINCHIP hs-cTnI) assay using plasma and whole blood from a healthy reference population. We also report analytical performance characteristics including values for LoB, LoD, LoQ, and within- and between-laboratory precision, and determine whether the assay meets accepted criteria for classification as a high-sensitivity test.

## METHODS AND MATERIALS

### SPINCHIP hs-cTnI ASSAY

The SPINCHIP hs-cTnI assay is a single-use fluorescence immunoassay device for quantitative measurement of cardiac troponin I (cTnI) in human finger-prick, Li-Heparin venous whole blood (“whole blood”) and Li-Heparin plasma (“plasma”) samples. No sample pre-treatment is necessary. Samples are collected from finger-prick or venous blood collection tubes using the cartridge-integrated sampler, with optional use of a sample transfer device for collection from closed tubes. The reagent cartridge is placed in the SPINCHIP Analyzer and processed. Upon initiation of the analysis, dual-axis centrifugal force separates blood cells from plasma and reconstitutes dried reagents within the cartridge. The cTnI antigen is captured on integrated antibody-coated beads in combination with fluorescently labelled antibodies. After washing, the fluorescent signal is measured. The SPINCHIP hs-cTnI assay result is displayed on the analyzer touchscreen, stored in memory, and, when connected, transferred to laboratory and hospital information systems.

### Analytical performance

Analytical performance of the SPINCHIP hs-cTnI assay was evaluated according to Clinical and Laboratory Standards Institute (CLSI) guidelines. Please refer to the Supplemental Methods section for a description of analytical studies involving limit of blank (LoB), LoD, limit of quantification (LoQ), linearity, overall imprecision, stability, batch-to-batch variation, interference, and cross-reactivity. Table S1 provides a summary of the parameters for LoB, LoD, LoQ, linearity, and precision studies.

### Precision at the 99^th^ percentile

Precision at the 99^th^ percentile was estimated using within-laboratory results for plasma and whole-blood samples obtained in two analytical performance studies. When combined, these studies comprised five SPINCHIP hs-cTnI cartridge batches and a total of 12 samples (cTnI range: 0.5–1968 ng/L) per sample type, corresponding to 28 independent precision estimates (4 samples×3 batches; 8 samples×2 batches). For plasma, precision was assessed over 20 days with two runs per day and two replicates per run, using one analyzer. Whole-blood samples were analyzed across four analyzers with ten replicates per analyzer on the day of collection. Imprecision was estimated independently for each sample-batch combination using ANOVA-based methods. For plasma samples, a two-factor nested ANOVA model (run nested within-day) was applied. For whole-blood samples, a one-factor ANOVA model with analyzer treated as a random effect was used. The objective was to determine whether the analytical imprecision at the 99^th^ percentile URL was <10% CV. Precision profiles were fitted using a three-parameter variance model, from which the precision at the 99^th^ URL was estimated according to: *σ*^2^=(*β*_0_ + *β*_1_*U*)*^Ϳ^*; where *σ*^2^=variance, *β*_0_=intercept, *β*_1_=change in variance with change in concentration (*U*), and *Ϳ* describes the curvature of the variance function. Samples with mean concentrations below the established LoQ values (plasma, 1.1 ng/L; whole blood, 1.4 ng/L) were excluded from precision modelling because their imprecision exceeds acceptance criteria and is not reliable in the clinically relevant range.

### Multi-site precision study

A multi-site, prospective, observational validation study assessed the precision of the SPINCHIP hs-cTnI assay in the hands of the intended users (could include nurses, laboratory staff, and research coordinators) utilizing plasma and whole blood. The study evaluated repeatability, within-laboratory precision, and reproducibility (total variation) for plasma samples, as well as repeatability for whole blood. Details are provided in the Supplemental Methods. This study was approved by the Norwegian Medical Products Agency (ID 24/21357-6; 2024-10-02) and the Norwegian Ethics Committees, Medical Devices (ID 698936; 2024-11-05).

### 99^th^ percentile study population

This was a multicenter, prospective, observational, non-randomized study designed to determine the 99th percentile URL for the SPINCHIP hs-cTnI assay, in accordance with IFCC Committee on Clinical Application of Biomarkers recommendations.(16) Individuals were healthy adults, approximately 50% male and 50% female, and evenly distributed across ages 18-80 years. This study was approved by the Swedish Medical Products Agency (ID 5.1.1-2025-026923; 2024-02-16).

Subjects were excluded if they had known cardiovascular disease, including history of AMI, angina, stroke, atrial fibrillation, peripheral vascular disease, deep vein thrombosis, pulmonary embolism, valvular disease, or heart failure. Individuals receiving treatment for hyperlipidemia or hypertension, or with investigator-assessed hypertension, were also excluded. Additional criteria included diabetes treatment (including diet-treated), body mass index <18 or >35 kg/m², and current smoking. Subjects with major illnesses or chronic conditions that could affect cardiac function, such as pulmonary, hepatic, or thyroid disease, or autoimmune disorders, were not eligible. A history of cancer within the preceding five years (except basal cell carcinoma *in situ*), acute hospitalization within the prior three months, or pregnancy also resulted in exclusion. Individuals previously enrolled were not permitted to participate again.

For inclusion, blood samples were evaluated for renal function, which had to be within acceptable limits, defined as an estimated glomerular filtration rate (eGFR) greater than 60 mL/min/1.73 m² for individuals aged 18–65 years and greater than 50 mL/min/1.73 m² for those older than 65 years. Subjects were also required to have a hemoglobin A1c (HbA1c) level below 6.5% (<48 mmol/mol) and either an N-terminal pro–B-type natriuretic peptide (NT-proBNP) concentration below 125 ng/L, or a B-type natriuretic peptide (BNP) concentration below 35 ng/L.

The determination of the 99^th^ percentile URLs was performed using a nonparametric approach to calculate the 99^th^ percentile of the hs-cTnI values, according to CLSI guideline EP28-A3C (19) and IFCC recommendations.(16) The per-protocol analysis set included all enrolled male and female participants who had at least one sample analyzed on the investigational device, provided they had no significant protocol deviations. The top 5% of hs-cTnI concentrations by cohort (female, male) and sample type (whole blood, plasma) were identified and corresponding frozen plasma samples were assessed to capture all samples potentially elevated due to human anti-mouse antibodies (HAMA) interference. The top 5% of values were chosen to capture all observations with the potential to substantially influence estimation of the 99^th^ URL. Samples with confirmed HAMA interference were excluded from the 99^th^ percentile analysis set but were retained in the LoD analysis set. The Dixon Q test was used to identify potential outliers.(20)

The LoD analysis set included all enrolled participants with at least one sample tested on the investigational device, with no significant protocol deviations, and results reported using an extended measurement range, capturing values below the LoD of the SPINCHIP hs-cTnI assay. This set was used to determine the proportion (%) of cTnI concentrations above the LoD. Of the 1070 subjects included in the 99^th^ percentile URL (n=1058) or LoD (n=436) sets, 424 subjects were included in both sets and 12 were included only in the LoD set.

### Sample-type validation

A single-center, prospective, observational, non-randomized sample-type comparison study was conducted to establish equivalence of hs-cTnI measurements from finger-prick capillary blood, plasma, and whole blood, using the SPINCHIP hs-cTnI assay, when performed by a healthcare professional. Hospitalized adult subjects, with available clinical cTn values, were screened for eligibility. Subjects were identified using a prescreening algorithm based on medical record cardiac troponin T results, ensuring coverage across the full analytical measuring range of SPINCHIP hs-cTnI (1.4–9000 ng/L). Clinically relevant concentrations, including the 99th percentile URL, rule-out, and rule-in thresholds, were deliberately enriched. Thirty-four (34) of the 150 planned samples were allocated to the low concentration range (1.4–30 ng/L). A total of 180 subjects were enrolled. Pregnant individuals, clinically unstable patients, and vulnerable populations (a compromised ability to either provide voluntary, informed consent or protect their own self-interest) were excluded. Duplicate measurements (replicates 1 and 2) were evaluated. Mean, %CV, SD, and mean difference were calculated; pooled %CV (mean squared %CV) and SD across the measuring range were also calculated. Mean difference across the measuring range was calculated with 95% CIs and 95% limits of agreement. This study was approved by the Norwegian Medical Products Agency (ID 23/29514-9; 2024-01-16) and the Norwegian Ethics Committees, Medical Devices (ID 698778; 2024-02-26).

All specimens used in clinical studies involving intended users were collected after informed consent and in accordance with protocols approved by the relevant Institutional Ethics Committees. All study procedures adhered to Good Clinical Practice and the ethical principles of the Declaration of Helsinki.

### Statistical analysis

Descriptive analyses, including the frequency and percentage of values above the LoD, were performed using SAS version 9.4 or later. The 99th percentiles and their corresponding confidence intervals were calculated using Analyze-it for Microsoft Excel, version 6.15.4 or later. Precision at the 99th percentile URL was evaluated separately using GraphPad Prism (version 11.0.1 or later).

## RESULTS

### Determination of the 99^th^ percentile URL

A total of 1355 subjects were screened for inclusion in the 99^th^ percentile URL study, of whom, 1100 were considered healthy and enrolled based on a health questionnaire and blood sampling to confirm normal HbA1c, eGFR and NT-proBNP/BNP. Following additional exclusions due to missing data (n=18 for plasma and n=16 for whole blood), the 55 samples (plasma and whole blood) with the highest cTnI concentration (top 5%) were evaluated for potential HAMA interference. Of these, 27 (∼50%) were excluded due to confirmed HAMA interference, resulting in per-protocol populations of 1055 and 1057 plasma and whole-blood samples, respectively (Figure S1). In plasma, 54.0% (n=570) were female, while the whole-blood population was 54.1% (n=572) females (Table S2, S3). For both plasma and whole-blood samples, ages spanned 18-80 years with mean of 45.5 (SD=15.8) years and median of 46.0 (Q1, Q3=31.0, 58.0) years. Participant age (18-80 years) was evenly distributed with representation across all decades. Ages were evenly distributed between sexes (Figure S2). Two paired samples were outliers with confirmed autoantibody/macrotroponin interference and were excluded in line with IFCC recommendations; this resulted in 1053 plasma and 1055 whole-blood samples used for analyses.(16)

The global 99^th^ percentile for plasma was 14.0 ng/L [95% CI: 10.6, 17.2] and for whole blood 13.7 ng/L [95% CI: 10.7, 18.0] (Table 1; Figure 1). Sex-specific 99^th^ percentiles for plasma were 15.7 ng/L [95% CI: 13.2, 24.9] for men and 10.3 ng/L [95% CI: 8.2, 14.7] for women.

**FIGURE 1.**
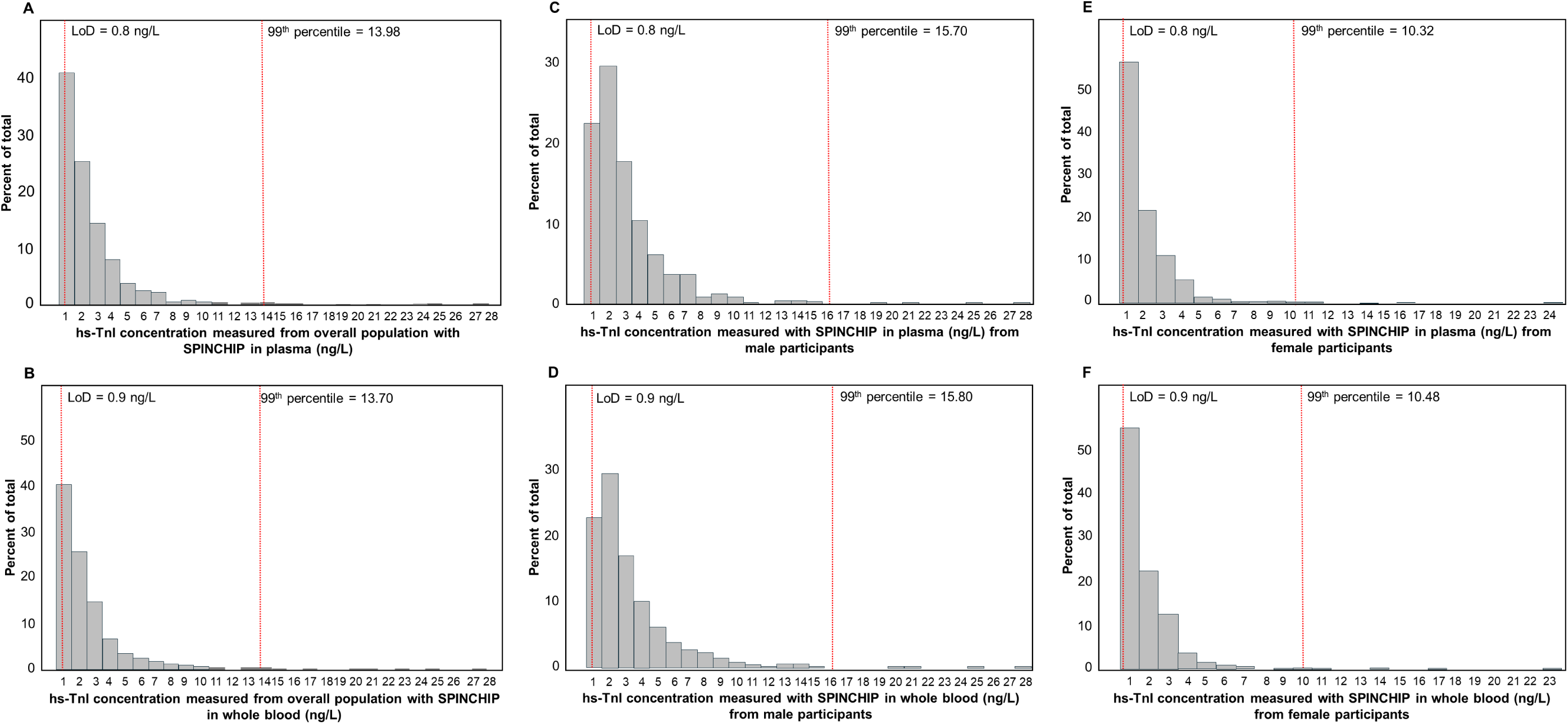
Distribution of cTnI values using SPINCHIP hs-cTnI in plasma samples from (A) the overall population (N=1053), (B) the male population (n=484), and (C) the female population (n=569). In whole-blood specimens the distributions are shown for (D) the overall population (N=1055), (E) the male population (n=484) and (F) the female population (n=571).

**Table 1.**
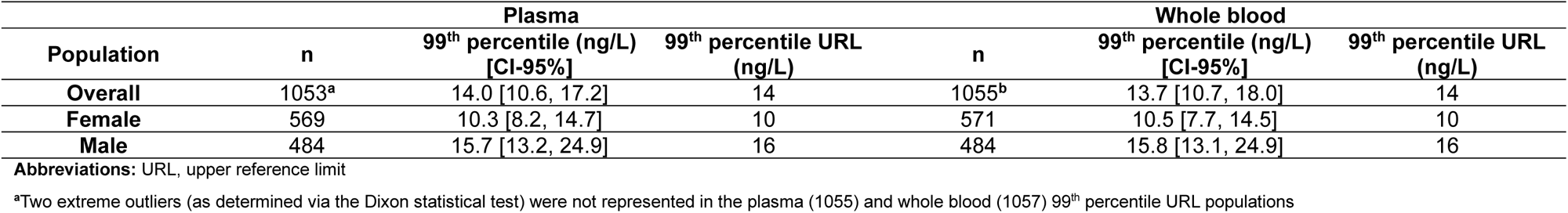
SPINCHIP hs-cTnI Determination of the 99^th^ Percentile URL.

### Percent above LoD

A total of 432 and 431 plasma and whole-blood samples, respectively, were analyzed to determine percent above LoD (Figure S1). The age distribution in the LoD population was balanced across sex and sample type, with a slight concentration in middle-aged subjects and no evidence of skewed distribution (Figure S3). Overall, 88.7% [95% CI: 85.3, 91.3] (383/432) and 85.2% [95% CI: 81.5, 88.2] (367/431) of plasma and whole-blood samples, respectively, were above the LoD. In both sample-type populations, the proportion of values above LoD was higher in males (≥94.0%) compared to females (≥76.1) (Table 2).

**Table 2.** cTnI values above the limit of detection^a^.

| Population | Total | Plasma |  | Total | Whole blood |  |
| --- | --- | --- | --- | --- | --- | --- |
|  |  | Values >LoD |  |  | Values >LoD |  |
|  |  | n | %, [CI-95%] |  | n | %, [CI-95%] |
| Overall | 432 | 383 | 88.7% [85.3, 91.3] | 431 | 367 | 85.2 [81.5, 88.2] |
| Female | 215 | 169 | 78.6% [72.6, 83.6] | 213 | 162 | 76.1 [69.9, 81.3] |
| Male | 217 | 214 | 98.6% [96.0, 99.7] | 218 | 205 | 94.0 [90.1, 96.5] |

**Abbreviations:** cTnI, cardiac troponin I; LoD, limit of detection
<sup>a</sup>Only subjects where the SPINCHIP hs-cTnI results were recorded using an extended results reporting range down to the LoD of the SPINCHIP hs-cTnI test could be used in the evaluation.

Across 3225 total measurements, performed by the intended users, the overall error rate was 3.5%. Most events were attributable to user-related issues rather than assay/instrument malfunction (Table S4). User errors, such as low or high sample volume, cartridge handling, or cartridge loading, accounted for ∼2.8% of runs. In contrast, non-user errors, including analyzer or cartridge related issues, were infrequent (0.7%).

### Analytical performance

High-sensitivity detection capability was demonstrated by a LoB of 0.3 ng/L in plasma and LoD of 0.8 ng/L for plasma and 0.9 ng/L for whole blood (Table S5). LoQ, defined as the lowest concentration quantified with a CV ≤20%, was 1.1 ng/L for plasma and 1.4 ng/L for whole blood (Table S6). The analytical measuring interval, defined as the range from LoQ to the upper limit of linearity, was demonstrated across a range from 1.1 ng/L to 9000 ng/L for the SPINCHIP hs-cTnI assay (Table S7).

Additional testing demonstrated that samples remained stable under the tested conditions (Table S8), showed shelf-life stability (Table S9) and stability during transport simulation (Tables S10, S11), with minimal batch-to-batch variability (Table S12). No Hook effect was observed for samples up to 1X10^6^ ng/L recombinant cTnI for SPINCHIP hs-cTnI (Table S13). Studies investigating HAMA and rheumatoid factor identified interference (≥10%) in 7/10 samples that included HAMA and in 6/10 samples that included rheumatoid factor (Table S14). Interference testing (Tables S15-S20) showed no meaningful effect from the substances evaluated (e.g., drug interference, thrombocytes, lipids), and cross-reactivity studies showed minimal signal contributions from the analytes tested (e.g., skeletal cTn, myoglobin) (Tables S21-24).

### Precision

Within-run (repeatability) and within-laboratory precision were evaluated across the analytical measurement range (0.7–10364 ng/L), with results summarized in Table S25. For plasma, repeatability (within-run) CVs ranged from 3.2% to 7.5% and for whole blood ranged from 3.2% to 8.0%. Within-laboratory CVs for plasma ranged from 3.7% to 7.7%. Both met predefined acceptance criteria, with CVs ≤20% for samples with cTnI concentrations ≤10 ng/L and ≤10% for samples with concentrations >10 ng/L.

Between-instrument reproducibility was demonstrated using 10 analyzers, 4 plasma samples, and over 5 days (Table S26). cTnI ranged from 10 to 4000 ng/L. The between-instrument CV ranged from 2% to 3% and overall CV ranged from 5% to 9%.

Within-laboratory precision was estimated at the 99^th^ percentile URL, with results presented in Table 3 and Figure 2. A total of 26 plasma and 25 whole-blood data points were included. For both matrices, imprecision was modelled using a three-parameter variance function. Based on the fitted precision profiles, the estimated imprecision at the respective 99th-percentile URLs was <10% CV for both plasma and whole blood. The CV at the global 99th percentile URL for both sample types was 5.8%. When stratified by sex-specific cut-offs, CV values at the 99^th^ URL were slightly higher in females (6.3–6.4%) and slightly lower in males (5.6%).

**FIGURE 2.**
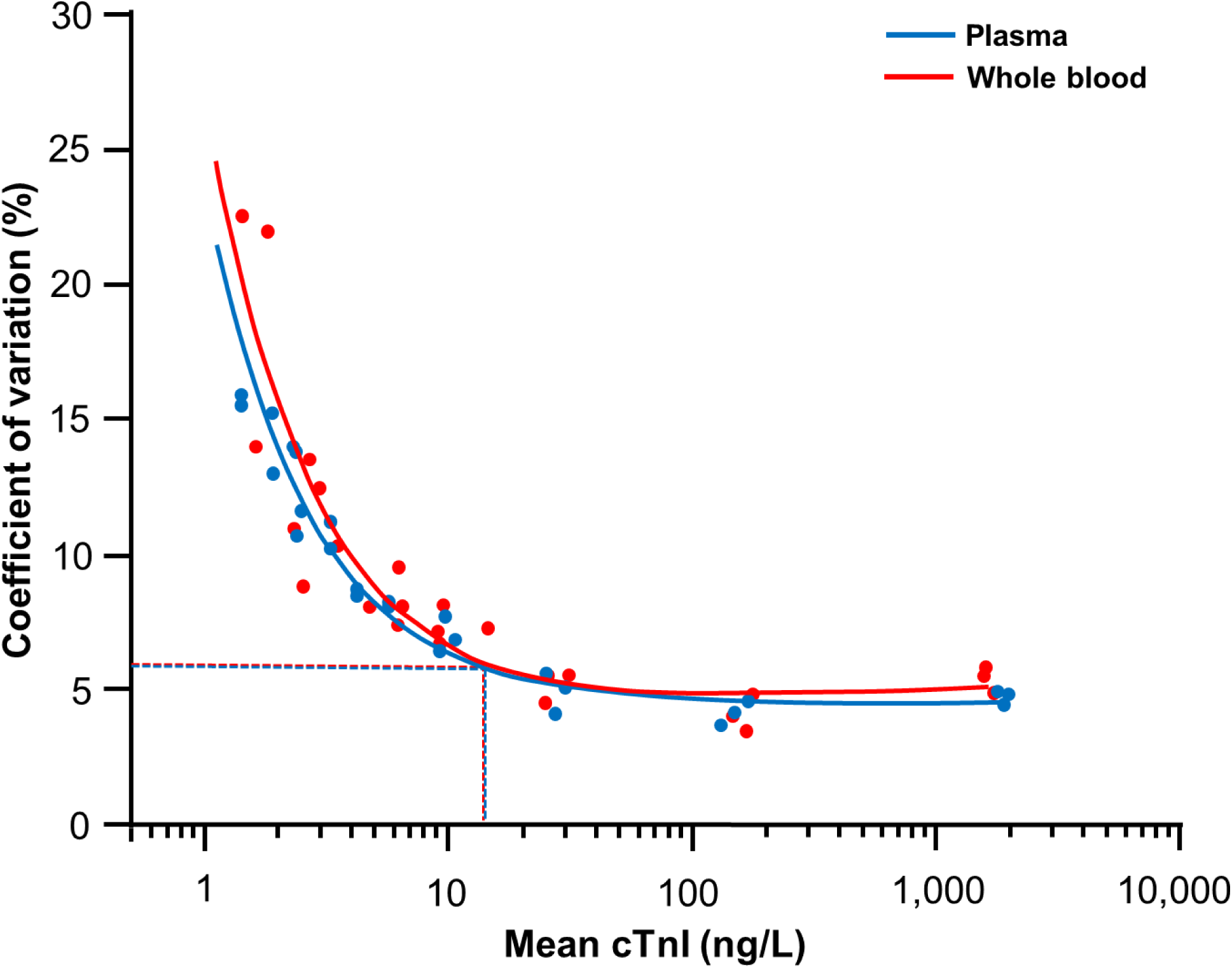
Three-parameter precision profiles for plasma (blue) and whole blood (red). Each point represents the mean concentration of a sample measured using a single cartridge batch. The dotted lines indicate the estimated %CV at the overall 99th-percentile upper reference limit for the SPINCHIP hs-cTnI assay (14 ng/L), derived from the variance function associated with the precision profiles. The variance function for plasma samples was: σ²=(0.1874 + 0.0446 U)^2.003^ and for whole blood was: σ²=(0.2376 + 0.0411 U)^2.079^

**Table 3.**
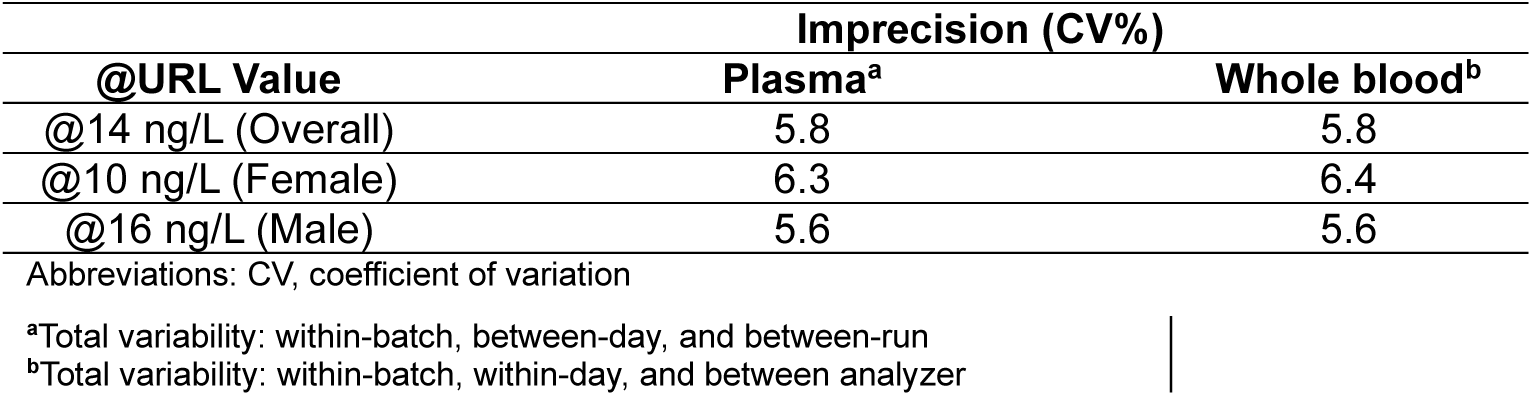
hs-cTnI precision estimation at the 99^th^ percentile.

| **Table 3. hs-cTnI precision estimation at the 99<sup>th</sup> percentile** |  |  |
| --- | --- | --- |
|  | **Imprecision (CV%)** |  |
| **@URL Value** | **Plasma<sup>a</sup>** | **Whole blood<sup>b</sup>** |
| @14 ng/L (Overall) | 5.8 | 5.8 |
| @10 ng/L (Female) | 6.3 | 6.4 |
| @16 ng/L (Male) | 5.6 | 5.6 |

Abbreviations: CV, coefficient of variation
<sup>a</sup>Total variability: within-batch, between-day, and between-run
<sup>b</sup>Total variability: within-batch, within-day, and between analyzer

Multi-site precision of the SPINCHIP hs-cTnI assay was evaluated in plasma and whole blood across multiple sites and a broad concentration range (∼3.6 to 7934.5 ng/L; Tables S27 and S28). In plasma, repeatability, within-laboratory precision, and between-site reproducibility showed comparable performance, with CVs ranging from 4.3% to 17.2%, and highest variability at the lowest concentration (∼3.7 ng/L). CVs were ≤8.5% at concentrations ≥19.8 ng/L. In whole blood, within-day repeatability CVs ranged from 3.3% to 13.9% across study sites, with the greatest variability observed at low concentrations (∼3.6–3.7 ng/L). Precision improved with increasing analyte concentration, with CVs ≤6.9% at ≥20 ng/L and ≤4.3% at higher concentrations (>2,000 ng/L). Overall, the assay demonstrated consistent precision across sample types, concentration ranges, and study sites.

### Sample-type validation

A total of 169 paired capillary and whole-blood samples, 169 paired capillary and plasma, and 170 paired plasma and whole-blood samples were included in a sample-type validation study. Across the full sample set, cTnI results obtained from capillary blood samples showed a mean difference of 4.5% versus whole blood and 2.1% versus plasma. For samples ≤20 ng/L, capillary blood samples showed a mean difference of 0.8 ng/L versus whole blood and 0.5 ng/L versus plasma. Plasma showed a mean difference of 2.6% versus whole blood across the full sample set and 0.4 ng/L in the ≤20 ng/L range (Table 4). Deming regression demonstrated a linear relationship between sample types (Figure S4).

**Table 4.**
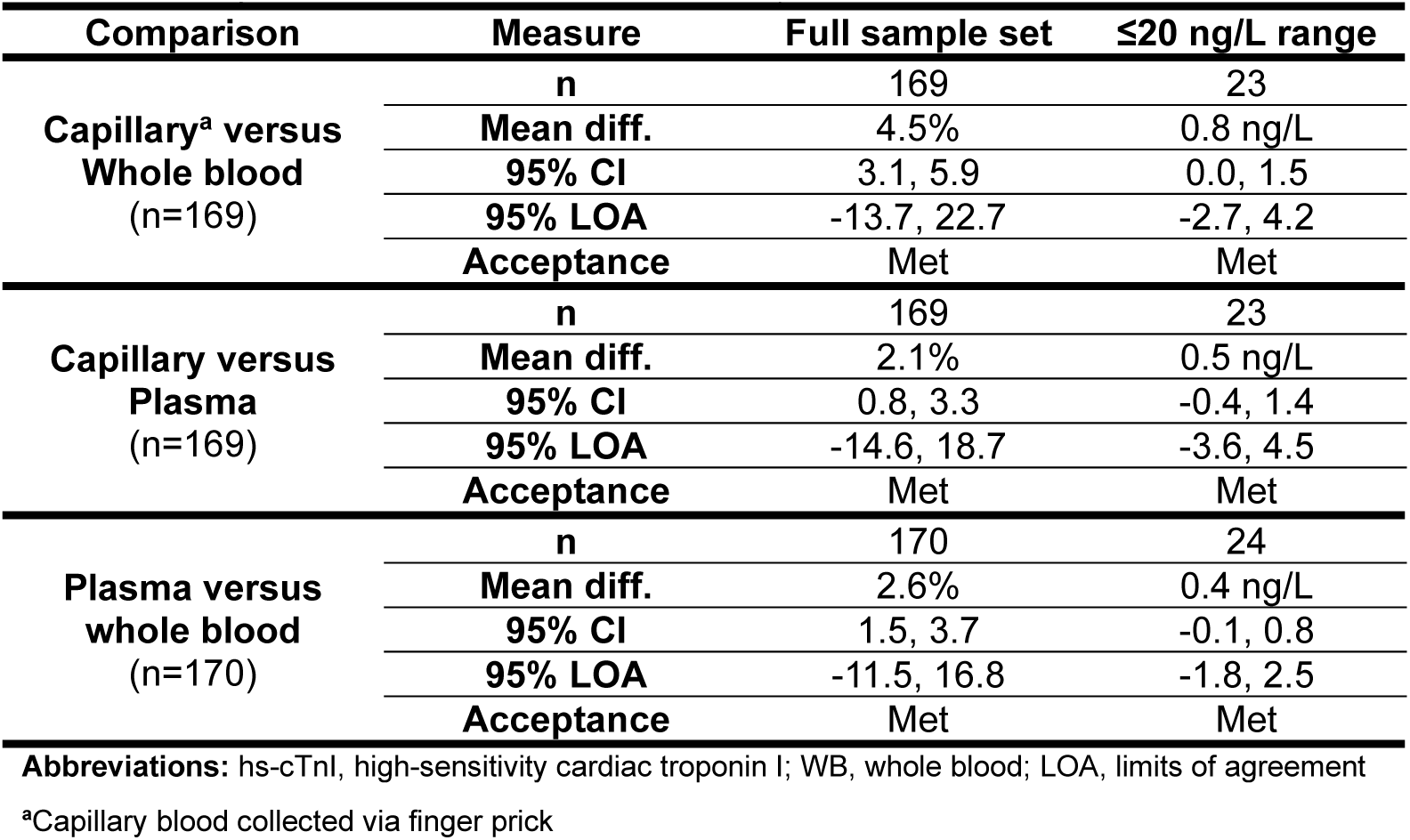
Sample-Type Validation: Capillary blood vs plasma vs whole blood in a clinical setting for the SPINCHIP hs-cTnI assay.

| Comparison | Measure | Full sample set | ≤20 ng/L range |
| --- | --- | --- | --- |
| **Capillary<sup>a</sup> versus Whole blood**<br>(n=169) | n | 169 | 23 |
|  | Mean diff. | 4.5% | 0.8 ng/L |
|  | 95% CI | 3.1, 5.9 | 0.0, 1.5 |
|  | 95% LOA | -13.7, 22.7 | -2.7, 4.2 |
|  | Acceptance | Met | Met |
| **Capillary versus Plasma**<br>(n=169) | n | 169 | 23 |
|  | Mean diff. | 2.1% | 0.5 ng/L |
|  | 95% CI | 0.8, 3.3 | -0.4, 1.4 |
|  | 95% LOA | -14.6, 18.7 | -3.6, 4.5 |
|  | Acceptance | Met | Met |
| **Plasma versus whole blood**<br>(n=170) | n | 170 | 24 |
|  | Mean diff. | 2.6% | 0.4 ng/L |
|  | 95% CI | 1.5, 3.7 | -0.1, 0.8 |
|  | 95% LOA | -11.5, 16.8 | -1.8, 2.5 |
|  | Acceptance | Met | Met |

**Abbreviations:** hs-cTnI, high-sensitivity cardiac troponin I; WB, whole blood; LOA, limits of agreement
<sup>a</sup>Capillary blood collected via finger prick

## DISCUSSION

This study evaluated the analytical performance of the SPINCHIP hs-cTnI assay and demonstrated robust precision when operated by intended users in a near-patient setting. Results involving LoB, LoD, and LoQ indicate high analytical sensitivity, allowing reliable detection and quantification of very low cTnI concentrations. The LoD and LoQ values reported here for SPINCHIP hs-cTnI should support the use of rule-out algorithms through reliable quantification of very low cTnI concentrations.(16–18, 21)

The overall 99^th^ percentile URL was 14 ng/L (sex-specific: 16 ng/L, men; 10 ng/L, women) for both plasma and whole blood. While concordant URLs across matrices do not alone establish analytical equivalence, they support consistent performance. This is further supported by sample-type comparison where plasma, whole blood, and capillary blood generated similar values in the hands of the intended user. Additionally, ≥94.0% of male and ≥76% of female cTnI values were above the LoD, satisfying IFCC criteria for high-sensitivity designation.(16, 17) Device-related errors were infrequent, and excluding user errors reduced the observed error rate from 3.5% to 0.7%, highlighting the importance of adequate operator training.

Imprecision studies across the measurement range, including the 99^th^ percentile URL, demonstrated CV% ≤10%, which met high-sensitivity analytical requirements.(18) The imprecision results were confirmed in this current prospective, multicenter analytical performance study, conducted by intended users under real-world conditions. This consistency across settings supports reproducible performance beyond controlled laboratory environments. Capillary sampling performed comparably to venous matrices, including at low concentrations critical for rule-out pathways. This supports capillary blood as a practical and reliable sample type, enhancing usability and clinical utility of SPINCHIP hs-cTnI point-of-care assay in settings where minimally invasive sampling provides operational advantages.

Preliminary analytical performance for SPINCHIP hs-cTnI has been previously reported.(22) The current study demonstrated comprehensive adherence to CLSI and IFCC guidance, covering detection capability, precision, linearity, interference, stability, and reproducibility. Inclusion of sample-type comparisons and multi-site precision assessments, conducted by intended users rather than research staff, reflects real-world performance in decentralized or point-of-care settings. Determination of the 99^th^ percentile URL used fresh samples, excluding those with documented biological interference (HAMA), which likely explains differences between the URL value reported here (14 ng/L) and that previously reported (32 ng/L).(22) Overall, these findings build on earlier work characterizing cTnI measurement by SPINCHIP hs-cTnI assay and confirm its robust analytical performance.

Recent evidence has demonstrated that antibody-mediated interference may disproportionately affect the upper tail of hs-cTn reference distributions and markedly increase calculated 99th percentile URLs.(16, 23) Here, the highest 5% of troponin values within the overall population were evaluated and those demonstrating HAMA interference were excluded from the analysis. The lower URL derived after exclusion of HAMA interference-positive samples may improve analytical sensitivity for myocardial injury detection; however, clinical diagnostic performance was not evaluated in this study.

Although HAMA interference may result in occasional false-positive cardiac troponin elevations,(24, 25) the clinical impact of such results is likely mitigated because AMI diagnosis requires integration of cTn findings with clinical evidence of myocardial ischemia, including symptoms, electrocardiographic changes, imaging findings, or identification of coronary thrombus.(14, 15) Furthermore, patients with elevated near-patient cTn results are typically evaluated using a central laboratory cTn assay during ongoing clinical assessment, which may help confirm or exclude misclassification due to interference during initial cTn testing. Consequently, the potential risk associated with rare HAMA-related false-positive results is likely outweighed by the benefit of a lower, interference-adjusted URL that reduces false-negative diagnoses and improves overall clinical sensitivity.

### Limitations

Although two statistical outliers were identified as positive for macrotroponin in this study, no systematic testing was employed to identify the presence of autoantibodies/macrotroponin in enrolled samples. Consequently, the ability of the assay to detect, differentiate, or mitigate the effects of autoantibodies/macrotroponin cannot be concluded from the data presented in this study. The enrollment population was predominantly (∼90%) of Northern-European descent. Findings may therefore not generalize to more diverse populations.

### Conclusions

SPINCHIP hs-cTnI demonstrated robust, reliable performance when operated by the intended user under real-world conditions, supporting routine clinical use. Overall, these results demonstrate SPINCHIP hs-cTnI meets the analytical performance requirements for hs-cTnI testing and is suitable for integration into AMI diagnostic algorithms.

## Data Availability

All data produced in the present study are available upon reasonable request to the authors

## A/AC/COI/F

## Acknowledgements

The authors would like to thank Tammy Bleak, PharmD, for her input and support during the development of this manuscript. The authors would also like to thank Ludovic Brossault and Didier Poirault for statistical support on this work.

## Author contributions

The authors contributed to this work as follows:

HR: Investigation; Methodology; Project administration; Supervision; Writing – original draft; Writing – review and editing.

JN: Investigation; Methodology (partial); Project administration; Writing – review and editing.

JM: Conceptualization; Data curation; Formal analysis; Investigation; Methodology; Project administration; Supervision; Validation; Writing – original draft; Writing – review and editing.

MNB: Conceptualization; Data curation; Formal analysis; Investigation; Methodology; Supervision; Validation; Writing – original draft; Writing – review and editing.

AO: Data curation; Formal analysis; Methodology; Validation; Visualization; Writing – review and editing.

DP: Conceptualization; Investigation; Methodology; Project administration; Resources; Supervision; Writing – review and editing.

SS (Stenmark): Conceptualization; Data curation; Investigation; Methodology; Software; Supervision; Writing – review and editing.

BBB: Conceptualization; Funding acquisition; Investigation; Project administration; Resources; Supervision; Writing – original draft; Writing – review and editing.

GS: Conceptualization; Data curation; Methodology; Project administration; Supervision; Validation; Writing – review and editing.

SS (Scott): Investigation; Methodology; Project administration; Supervision; Writing – review and editing.

SM-B: Formal analysis; Investigation; Methodology; Writing – review and editing. LO: Writing – review and editing.

MNL: Investigation; Methodology; Project administration; Supervision; Writing – review and editing.

AT: Investigation; Methodology; Project administration; Supervision; Validation; Writing – review and editing.

KA: Conceptualization; Investigation; Methodology; Supervision; Visualization; Writing – original draft; Writing – review and editing.

## Notes

The study protocol related to this work can be obtained via communication with the corresponding author

