## Supplemental Information for "Analytical Performance and 99th Percentile Upper Reference Limit of the Novel SPINCHIP High-Sensitivity Cardiac Troponin I Point-of-Care Assay"

*Author Byline*

Jenny MacKenzie, PhD,[1] Kristin Moberg Aakre, MD, PhD,[2] Didrik Paus, PhD,[1] Marianne Nordlund Broughton, PhD,[1] Gro Leite Storvold, PhD,[1] Anniken Olberg, MSc,[1] Sebastian Stenmark,[1] Birgitte Boonstra Booij, PhD,[1] Sara Scott,[1] Sandrine Michel-Busseret, PhD,[3] Lydia Octave, PhD,[4] Arnljot Tveit, MD, PhD,[5] Magnus N. Lyngbakken, MD, PhD,[6,7] Johan Nilsson, MD, PhD,[8] Helge Røsjø, MD, PhD,[6,7*]

*Author affiliations*

[1]bioMérieux Norge, Hoffsveien 21/23, 0275 Oslo, Norway
[2]Haukeland University Hospital and Institute of Clinical Science, University of Bergen, Jonas Lies vei 65, 5021 Bergen, Norway
[3]bioMérieux SA, 376 Chemin de l'Orme, 69280 Marcy-l'Etoile, France
[4]bioMérieux Inc., 100 Rodolphe Street, Durham, NC, 27712
[5]Bærum Hospital, Sogneprest Munthe-Kaas vei 100, 1346 Gjettum, Norway
[6]Akershus Clinical Research Center (ACR), Division of Research and Innovation, Akershus University Hospital, Lørenskog, Norway
[7]K. G. Jebsen Center for Cardiac Biomarkers, Institute for Clinical Medicine, University of Oslo, Oslo, Norway
[8]Clinical Trial Consultants AB (CTC), Dag Hammarskjölds väg 10B, 752 37 Uppsala, Sweden

*Corresponding author:*

Helge Røsjø, MD, PhD
Akershus University Hospital
Sykehusveien 25, 1478 Lørenskog, Norway


SUPPLEMENTAL MATERIAL

### SUPPLEMENTAL METHODS

#### Analytical Outcomes

*Detection capability*

The limit of blank (LoB; highest cTnI value expected when testing samples that do not contain any cTnI); limit of detection (LoD; lowest cTnI value reliably detected above normal background noise), and limit of quantitation (LoQ; concentration at which quantitative results are considered acceptable for clinical use) for the SPINCHIP hs-cTnI assay were established in accordance with CLSI EP17-A2.(1) The LoB study was performed by three operators across three days and LoB data were generated using cTn–depleted plasma. A total of 240 measurements were obtained, 60 measurements per sample were obtained for each of four SPINCHIP hs-cTnI cartridge batches (Table S1). The LoB was calculated using a nonparametric approach; the 95th-percentile rank was determined by the equation: $Rank_{95}=0.5+(0.95\times B)$, with $B$ representing the total number of data points (n=240). The cTnI concentration residing at the 95th-percentile rank was recorded as the LoB.

The LoD plasma study was performed over three days and included four samples spanning 1-5 times the LoB (0.3 to 1.5 ng/L) (Table S1). LoD from whole blood also included four samples, spanning the same range as with plasma; however, LoD testing on whole blood occurred over one day as frozen storage of whole blood precludes sample validity. The LoD was estimated using the parametric method in CLSI EP17-A2,(1) based on all low-level sample measurements obtained across the four cartridge batches. The LoD was calculated from the pooled standard deviation of the combined data ($SD_L$), the LoB, and the false negative threshold (β=0.05) for the 95th percentile of a normal distribution ($cp$) according to the following equation: LoD = LoB +

*cp* X $SD_L$. The LoB obtained for plasma samples was utilized for LoD calculation on whole blood samples.

The objective for the LoQ study was to determine the lowest level of cTnI concentration that can be quantified by the SPINCHIP hs-cTnI assay with a within-laboratory CV of ≤ 20%. LoQ data for SPINCHIP hs-cTnI were determined using plasma (9 samples with 1440 total measurements) and whole blood (8 samples with 640 total measurements) across a cTnI concentration range of 1.1 to 5.0 ng/L (Table S1).

*Linearity*

The linearity study for the SPINCHIP hs-cTnI assay followed CLSI EP06-Ed2 guidelines and used an eleven-level dilution panel prepared from a high-cTnI plasma pool and a cTnI buffer matrix, the latter selected to avoid negative bias observed with depleted plasma.(2) Three cartridge batches were tested, with 7–14 replicates per dilution level depending on repeatability requirements (CV ≤10% and allowable bias [the difference between the observed or measured result and the value predicted by the linear model] of 10%). Seven SPINCHIP analyzers were used per level, each generating one replicate per sample per analyzer, and the entire panel for each batch was run in a single day. Linearity assessment employed weighted linear regression, with integrity checks and outlier evaluation performed according to EP06.(2) Dilution samples spanning <1.1 to >9000 ng/L were prepared gravimetrically, corrected for density, aliquoted, frozen, and analyzed to confirm assay linearity across the measurement range.

*Precision studies*

Imprecision at the 99$^{th}$ percentile was determined from repeatability (within-run) and within-laboratory precision analysis based on results from SPINCHIP hs-cTnI using plasma and whole blood (repeatability only) samples. Testing for plasma (960 total replicates) and whole blood (480 total replicates) samples was performed using 4 samples across three SPINCHIP hs-cTnI cartridge batches (Table S1). The target cTnI concentrations for samples one through four were 10 ± 5 ng/L, 30 ± 10 ng/L, 200 ± 40 ng/L, and 2000 ± 50 ng/L, respectively. The objective was to determine whether the repeatability and overall imprecision met the following requirements: cTnI ≤10 ng/L with a CV ≤20% or a cTnI concentration of >10 ng/L with a CV ≤10%.(1) Plasma samples included two replicates, using three cartridge batches, across two runs, over 20 days. For whole blood, ten replicates were included per sample, using three cartridge batches, across four different analyzers. For each plasma sample, the mean concentration, within-series precision, and total precision (reflecting variability between series, days, and instruments) were calculated using a nested ANOVA model incorporating instrument, day, run, and repeatability effects. Both repeatability and total imprecision were reported as SD and CV. Only repeatability was calculated for whole blood as testing over multiple days was not possible for whole blood due to preclusion by freeze-thaw.

*Multi-instrument precision study*

A study was performed using SPINCHIP hs-cTnI to determine reproducibility across instruments. The study was conducted over a seven-day period by four trained operators; with 1–2 operators performing testing each day. Two fresh and two frozen plasma samples, obtained from healthy donors under informed consent, were used to prepare the four final study samples: one native (healthy), and three spiked with patient plasma pools or recombinant cardiac troponin to achieve the targeted cTnI concentration ranges (2 to 10 ng/L; 20 to 50 ng/L; 100 to 500 ng/L; ≥2000 ng/L; Table S1). A total of ten SPINCHIP analyzers were used, and each sample was analyzed in five replicates per analyzer on each of the five testing days, yielding 25 replicates per instrument and 250 replicates per sample. Error runs were repeated on the same instrument within the same day. Reproducibility and between-instrument precision were assessed using a two-factor nested ANOVA model.

*Multi-site precision study*

A multi-site, prospective, observational, and non-randomized validation study was conducted to establish the within-day (repeatability), within-laboratory (imprecision), and overall reproducibility (total analytical variation) of the SPINCHIP hs-cTnI assay, in the hands of the intended user, when measured on plasma; and to determine repeatability with whole blood. The study enrolled and tested samples from adult subjects with known cardiac troponin concentrations from two clinical sites (sites 1 and 2); the third site functioned solely as a testing laboratory. All participants were screened using a finger-prick sample analyzed with the SPINCHIP hs-cTnI assay to identify individuals falling within predefined concentration ranges. Samples selected for precision evaluation were chosen to span the expected 99th-percentile cut-off, key clinical decision thresholds (rule-out and rule-in); and low, medium, and high concentrations across the assay's measuring range (1.1–9000 ng/L). In total, 37 subjects were screened, of whom ten met the target concentration criteria. Five subjects provided samples for plasma precision testing at all three sites and whole blood repeatability testing at sites one and two, separately; five additional subjects contributed samples for whole blood repeatability testing at site three. For plasma, the five samples were run in replicates of five, across five days and across the three testing sites (375 total measurements), whereas for whole blood, five samples were run in replicates of ten, across one day, with four analyzers, across three testing sites (600 total measurements) (Table S1).

*Stability*

Real-time stability of SPINCHIP hs-cTnI cartridges was evaluated in accordance with CLSI EP25A.(3) The stability (the ability of an assay, its reagents, or its measured analyte to maintain performance characteristics within predefined acceptance criteria over a specified period of time under defined storage or operating conditions) studies included a total of six cartridge batches, comprising two shelf-life (defined as the period of time during which an assay [reagents, cartridges, calibrators, etc.] remains fit for use and performance remains within specification when stored under the manufacturer's specified conditions) studies (3 batches per study) and two simulated transport studies (one batch per study). The cartridges were stored under recommended storage conditions (2–8 °C) and at room temperature (23-27 °C), with testing performed at predefined intervals. Stability at 23-27 °C was also verified at the end of shelf life by transferring a subset of cartridges to room temperature after 9 months of refrigerated storage. Analytical performance was assessed using plasma samples spanning the SPINCHIP hs-cTnI measuring range. Each sample was analyzed in 10–20 replicates per time point, using ten SPINCHIP analyzers. For the shelf-life evaluation, drift from baseline was estimated from linear regression of mean cTnI results obtained at each time point. For the transport studies, cartridges were exposed to a sequence of physical and/or environmental stress conditions prior to storage at the abovementioned temperatures. The obtained cTnI results at each test point were compared to unstressed cartridges of the same batch and storage condition.

*Batch-to-batch variation*

Batch-to-batch variation of the SPINCHIP hs-cTnI assay was evaluated using 74 plasma samples spanning a concentration range of 3 to 5000 ng/L. Each sample was analyzed in a single replicate across three cartridge batches (total of three measurements per sample), using the same analyzer for all replicates of a given sample. If replicate differences exceeded predefined criteria based on assay precision, additional measurements were performed. Batch-to-batch variation was assessed by pairwise comparison of results between cartridge batches using Bland–Altman analysis and Passing–Bablok regression. Bias was evaluated across predefined concentration intervals, with separate analyses performed for low and higher concentration ranges due to mixed variability.

*Hook effect*

The potential for a high-dose hook effect was evaluated using plasma samples spiked with recombinant cardiac troponin ITC complex across a concentration range of 100 to $1x10^6$ ng/L, as well as native patient plasma samples with high cTnI concentrations (~60600 to ~403660 ng/L). Recombinant samples were prepared by serial dilution of ITC stock solutions into plasma, while patient samples were obtained commercially and pre-characterized using an external assay. All samples were analyzed in four replicates (one per analyzer) using the SPINCHIP hs-cTnI system. Hook effect was assessed by plotting raw signal versus cardiac troponin concentration and evaluating for any decline in signal at increasing concentrations; results for samples exceeding the measuring range were reported as ≥9000 ng/L.

#### Interference and Cross-Reactivity Studies

*Heterophilic Antibody Interference*

Interference from heterophilic antibodies was evaluated using serum samples containing human anti-mouse antibodies (HAMA; n = 12) or rheumatoid factor (RF; n = 13), together with control samples from healthy individuals (n = 5). Samples containing elevated levels of HAMA or RF were spiked with a cardiac troponin-rich plasma pool to achieve target cTnI concentrations of approximately 30–100 ng/L (target ~50 ng/L), while control samples were spiked with an equal volume of the same pool. All samples were analyzed in nine replicates across multiple SPINCHIP analyzers following verification of assay performance using control materials. Interference was assessed by comparing cTnI responses in HAMA- or RF-positive samples relative to the mean response of spiked control samples and expressed as percent difference.

*Exogenous compounds (drugs)*

Interference from pharmaceutical compounds was evaluated according to CLSI EP07 and EP37 guidelines using pooled plasma samples at two cTnI levels (~15–60 ng/L and ~500–1500 ng/L).(4, 5) Samples were spiked with individual compounds at clinically relevant concentrations, while matched controls were prepared using corresponding solvents. All samples were analyzed in nine replicates across multiple SPINCHIP analyzers following verification with control materials. If assay imprecision exceeded predefined criteria, replicate numbers were increased. Interference was assessed as the percent difference between drug-spiked samples and controls. For compounds exceeding acceptance limits, additional dose–response testing was performed at reduced concentrations.

*Endogenous interferents (bilirubin, hemoglobin, lipids)*

Interference from conjugated and unconjugated bilirubin, hemoglobin, and lipids (a mixture of neutral triglycerides and predominantly unsaturated fatty acids) was evaluated in accordance with CLSI EP07 and EP37 guidelines using pooled plasma samples at low and high cTnI concentrations (~15–60 ng/L and ~500–1500 ng/L).(4, 5) Samples were spiked with each substance at clinically relevant concentrations, while matched controls were prepared with corresponding solvents. Lipid interference was assessed using Intralipid (20% emulsion) at 2000 mg/dL. All samples were analyzed in nine replicates across multiple analyzers following validation with control materials, with additional replicates performed if required. Interference was calculated as the percentage difference between spiked samples and their respective controls.

*Thrombocytes and leukocytes*

Interference from blood cells was assessed using whole blood samples with elevated thrombocyte (300–500 $\times 10^9$/L; n = 10) or leukocyte (>10 $\times 10^9$/L; n = 6) levels. For thrombocyte studies, both native and cTnI-spiked samples were included, while leukocyte studies used samples with native measurable cTnI concentrations. Corresponding plasma controls were prepared from each sample by centrifugation to remove cells. Whole blood and matched plasma samples were analyzed in nine replicates across multiple analyzers. Interference was determined by comparing cTnI concentrations in cell-rich whole blood to paired plasma controls and expressed as percent difference or absolute bias.

*Hematocrit*

Interference from elevated hematocrit was evaluated using whole blood samples manipulated to achieve hematocrit levels of approximately 43–64%. Samples were spiked with recombinant cTnI (~50–200 ng/L) prior to adjustment. Corresponding plasma samples served as controls. Test and control samples were analyzed in nine replicates across multiple analyzers following validation with control materials. Interference was assessed by comparing cTnI results across hematocrit levels relative to controls and expressed as percent difference and recovery.

*Cross-Reactivity Studies*

Cross-reactivity was evaluated in accordance with CLSI EP07 guidelines using plasma samples at three cTnI levels: depleted (<1.1 ng/L), medium (~15–60 ng/L), and high (~500–1500 ng/L).(4) For each analyte, test samples were spiked with the potential cross-reactant at $1X10^6$ ng/L, while matched controls received an equivalent volume of control solution without the cross-reactant. All samples were analyzed in nine replicates across multiple SPINCHIP analyzers and across three cartridge batches following verification with control materials. The evaluated cross-reactants included skeletal troponin I (skTnI), cardiac troponin C (cTnC), and cardiac troponin T (cTnT), creatine kinase myocardial band (CK-MB), and myoglobin. Cross-reactivity was calculated as the difference in measured cTnI concentration between spiked and control samples, normalized to the concentration of the added cross-reactant, and expressed as a percentage.

#### REFERENCES

1. Clinical and Laboratory Standards Institute. 2012. EP17-A2: Evaluation of Detection Capability for Clinical Laboratory Measurement Procedures. 2nd ed. Wayne, PA: CLSI.

2. Clinical and Laboratory Standards Institute (CLSI). Evaluation of the Linearity of Quantitative Measurement Procedures; Approved Guideline—Second Edition. CLSI document EP06-A2. Wayne, PA: CLSI; 2003.

3. Clinical and Laboratory Standards Institute (CLSI). Evaluation of Stability of In Vitro Diagnostic Reagents; Approved Guideline. CLSI document EP25-A. Wayne, PA: CLSI; 2009.

4. Clinical and Laboratory Standards Institute (CLSI). Interference Testing in Clinical Chemistry; Approved Guideline—Second Edition. CLSI document EP07-A2. Wayne, PA: CLSI; 2005.

5. Clinical and Laboratory Standards Institute (CLSI). Supplemental Tables for Interference Testing in Clinical Chemistry. CLSI document EP37. Wayne, PA: CLSI; 2018.

**SUPPLEMENTAL TABLES**
**TABLE S1**

**Table S1.** Study parameters for analytical testing during validation of SPINCHIP hs-cTnI

| Analytical characteristic | Matrix | Analyte concentration | Number | | | | | | | |
|---|---|---|---|---|---|---|---|---|---|---|
| | | | Samples | Replicates | Cartridge batch | Analyzers | Days (d) or time points | Runs | Sites | Total |
| LoB (Table S5) | Plasma | Blank | 4 | 1 | 4 | 5 | 3 d | 1 | 1 | 240 |
| LoD (Table S5) | Plasma | Very low[a] | 4 | 2 | 4 | 4 | 3 d | 1 | 1 | 384 |
| | WB[b] | | 4 | 6 | 4 | 4 | 1 d | 1 | 1 | 384 |
| LoQ (Table S6) | Plasma | Low[c] | 9 | 2 | 2 | 1 | 20 d | 2 | 1 | 1440 |
| | WB[b] | | 8 | 10 | 2 | 4 | 1 d | 1 | 1 | 640 |
| Linearity (Table S7) | Plasma | Very low to high[d] | 11 | 1 or 2[e] | 3 | 7 | 1 d | 1 | 1 | 308 |
| Within-lab (Table S25) | Plasma | Low to high | 4 | 2 | 3 | 1 | 20 d | 2 | 1 | 960 |
| Repeatability (Table S25) | WB | | 4 | 10 | 3 | 4 | 1 d | 1 | 1 | 480 |
| Between-instrument (Table S26) | Plasma | Low to high[f] | 4 | 5 | 1 | 10 | 5 d | 1 | 1 | 1000 |
| Multi-site (Table S27 & S28) | Plasma | Low to high[g] | 5 | 5 | 1 | 1 | 5 | 1 | 3 | 375 |
| | WB[b] | | 5 | 10 | 1 | 4 | 1 | 1 | 3 | 600 |

**Abbreviations:** cTnI, cardiac troponin I; LoB, limit of blank; LoD, limit of detection; WB, whole blood; LoQ, limit of quantification

[a]Very low level samples contained cTnI concentrations that were 1-5 times the LoB (0.3 to 1.5 ng/L)
[b]Multiple day testing not possible for whole blood due to a preclusion by freeze-thaw
[c]Low concentrations from 1.1 to 5.0 ng/L; 5 ng/L is the highest acceptable concentration for limit of quantitation
[d]Very low to high concentrations spanned a range from 0.7 to 10364 ng/L
[e]Study was run with one replicate per analyzer; if the repeatability was outside the validity criterion (coefficient of variation ≤10%), a second replicate was run across the seven analyzers to account for higher imprecision in the cTnI range
[f]Low to high concentrations: sample 1 = 2 to 10 ng/L; sample 2 = 20 to 50 ng/L; sample 3 = 100 to 500 ng/L; sample 4 = >2000 ng/L
[g]Low to high target concentrations were: 5 to 12 ng/L; 20 to 40 ng/L; 100 to 400 ng/L; 1000 to 3000 ng/L; 4500 to 9000 ng/L

**TABLE S2**

**Table S2.** Demographics and baseline characteristics of subjects providing plasma and whole blood; per protocol set for the 99th URL study

| | | Plasma | | | Whole blood | | |
|---|---|---|---|---|---|---|---|
| **Demographic** | **Statistic** | **Female** (n=570) | **Male** (n=485) | **All** (n=1055) | **Female** (n=572) | **Male** (n=485) | **All** (n=1057) |
| **Age** (years) | n (missing values) | 570 (0) | 485 (0) | 1055 (0) | 572 (0) | 485 (0) | 1057 (0) |
| | Mean (SD) | 45.1 (16.1) | 46.0 (15.5) | 45.5 (15.8) | 45.1 (16.1) | 46.0 (15.5) | 45.5 (15.8) |
| | Median [Q1; Q3] | 45 [30; 57] | 48 [32; 58] | 46 [31; 58] | 45 [29.5; 57] | 48 [32; 58] | 46 [31; 58] |
| | [Min; Max] | [18; 80] | [18 80] | [18; 80] | [18; 80] | [18; 80] | [18; 80] |
| **Ethnicity** | N (Missing values) | 570 (0) | 485 (0) | 1055 (0) | 572 (0) | 485 (0) | 1057 (0) |
| | Northern European | 389 (68.2%) | 296 (61.0%) | 685 (64.9%) | 391 (68.4%) | 296 (61.0%) | 687 (65.0%) |
| | Other European | 121 (21.2%) | 123 (25.4%) | 244 (23.1%) | 121 (21.2%) | 123 (25.4%) | 244 (23.1%) |
| | Other | 60 (10.5%) | 66 (13.6%) | 126 (11.9%) | 60 (10.5%) | 66 (13.6%) | 126 (11.9%) |
| **Height** (cm) | n (missing values) | 570 (0) | 485 (0) | 1055 (0) | 572 (0) | 485 (0) | 1057 (0) |
| | Mean (SD) | 166.2 (6.6) | 179.8 (6.8) | 172.5 (9.5) | 166.2 (6.6) | 179.8 (6.8) | 172.5 (9.5) |
| | Median [Q1; Q3] | 166 [162; 171] | 180 [175; 184] | 172 [165; 180] | 166 [162; 171] | 180 [175; 184] | 172 [165; 180] |
| | [Min; Max] | [147; 187] | [158; 210] | [147; 210] | [147; 187] | [158; 210] | [147; 210] |
| **Weight** (kg) | n (missing values) | 570 (0) | 485 (0) | 1055 (0) | 572 (0) | 485 (0) | 1057 (0) |
| | Mean (SD) | 68.3 (10.7) | 82.8 (11.7) | 75.0 (13.3) | 68.3 (10.7) | 82.8 (11.7) | 75.0 (13.3) |
| | Median [Q1; Q3] | 67 [61; 74.7] | 81.9 [74.8; 90.4] | 73.9 [64.8; 83.8] | 67 [61; 74.6] | 81.9 [74.8; 90.4] | 73.9 [64.8; 83.8] |
| | [Min; Max] | [43.1; 109.5] | [53.7; 123.6] | [43.1; 123.6] | [43.1; 109.5] | [53.7; 123.6] | [43.1; 123.6] |
| **Pulse rate** (bpm) | n (missing values) | 570 (0) | 485 (0) | 1055 (0) | 572 (0) | 485 (0) | 1057 (0) |
| | Mean (SD) | 70.7 (12.5) | 68.4 (12.0) | 69.6 (12.3) | 70.7 (12.5) | 68.4 (12) | 69.6 (12.3) |
| | Median [Q1; Q3] | 69 [62; 77] | 67 [60; 76] | 69 [61; 77] | 69 [62; 77] | 67 [60; 76] | 69 [61; 77] |
| | [Min; Max] | [40; 141] | [40; 109] | [40; 141] | [40; 141] | [40; 109] | [40; 141] |
| **Diastolic bp** (mmHg) | n (missing values) | 570 (0) | 485 (0) | 1055 (0) | 572 (0) | 485 (0) | 1057 (0) |
| | Mean (SD) | 76.6 (8.4) | 79.2 (8.4) | 77.8 (8.5) | 76.6 (8.4) | 79.2 (8.4) | 77.8 (8.5) |
| | Median [Q1; Q3] | 76 [71; 83] | 80 [73; 86] | 78 [72; 84] | 76 [71; 83] | 80 [73; 86] | 78 [72; 84] |
| | [Min; Max] | [50; 99] | [51; 105] | [50; 105] | [50; 99] | [51; 105] | [50; 105] |
| **Systolic bp** (mm Hg) | n (missing values) | 570 (0) | 485 (0) | 1055 (0) | 572 (0) | 485 (0) | 1057 (0) |
| | Mean (SD) | 118.1 (12.9) | 128.5 (10.9) | 122.9 (13.1) | 118.1 (12.9) | 128.5 (10.9) | 122.9 (13.1) |
| | Median [Q1; Q3] | 116.5 [109; 128] | 129 [121; 137] | 123 [114; 133] | 116.5 [109; 128] | 129 [121; 137] | 123 [114; 133] |
| | [Min; Max] | [76; 166] | [90; 165] | [76; 166] | [76; 166] | [90; 165] | [76; 166] |
| **At least one concomitant medication; n (%)** | | 422 (74.0%) | 245 (50.5%) | 667 (63.2%) | 425 (74.3%) | 245 (50.5%) | 670 (63.4%) |

TABLE S3

**Table S3.** Concomitant medications for subjects providing plasma and whole blood; per protocol sets

| | Plasma | | | Whole blood | | |
|---|---|---|---|---|---|---|
| **Medications** | **Female** (n=570) | **Male** (n=485) | **All** (n=1055) | **Female** (n=572) | **Male** (n=485) | **All** (n=1057) |
| At least one | 422 (74.0%) | 245 (50.5%) | 667 (63.2%) | 425 (74.3%) | 245 (50.5%) | 670 (63.4%) |
| Analgesics, muscle relaxants | 93 (16.3%) | 40 (8.2%) | 133 (12.6%) | 93 (16.3%) | 40 (8.2%) | 133 (12.6%) |
| Antibacterials, antifungals, Antimycotics, Antiprozoals | 5 (0.9%) | 2 (0.4%) | 7 (0.7%) | 5 (0.9%) | 2 (0.4%) | 7 (0.7%) |
| Antidiarrheals, intestinal anti-inflammatory/anti-infective agents | 15 (2.6%) | 8 (1.6%) | 23 (2.2%) | 15 (2.6%) | 8 (1.6%) | 23 (2.2%) |
| Anti-inflammatory and antirheumatic products | 100 (17.5%) | 49 (10.1%) | 149 (14.1%) | 101 (17.7%) | 49 (10.1%) | 150 (14.2%) |
| Antithrombotic agents | 2 (0.4%) | 0 (0.0%) | 2 (0.2%) | 2 (0.4%) | 0 (0.0%) | 2 (0.2%) |
| Antiviral agents | 1 (0.2%) | 3 (0.6%) | 4 (0.4%) | 1 (0.2%) | 3 (0.6%) | 4 (0.4%) |
| Beta blocking agents (for treatment of essential tremor and palpition) | 2 (0.4%) | 1 (0.2%) | 3 (0.3%) | 2 (0.4%) | 1 (0.2%) | 3 (0.3%) |
| Corticosteroids | 3 (0.5%) | 7 (1.4%) | 10 (0.9%) | 3 (0.5%) | 7 (1.4%) | 10 (0.9%) |
| Dermatologicals | 3 (0.5%) | 5 (1.0%) | 8 (0.8%) | 3 (0.5%) | 5 (1.0%) | 8 (0.8%) |
| Dietary supplements | 190 (33.3%) | 92 (19.0%) | 282 (26.7%) | 191 (33.4%) | 92 (19.0%) | 283 (26.8%) |
| Drugs for acid related disorders; constipation | 37 (6.5%) | 24 (4.9%) | 61 (5.8%) | 37 (6.5%) | 24 (4.9%) | 61 (5.8%) |
| Drugs for obstructive airway diseases | 13 (2.3%) | 16 (3.3%) | 29 (2.7%) | 14 (2.4%) | 16 (3.3%) | 30 (2.8%) |
| Drugs for treatment of osteoporosis | 2 (0.4%) | 0 (0.0%) | 2 (0.2%) | 2 (0.4%) | 0 (0.0%) | 2 (0.2%) |
| Drugs used in diabetes (used for weight loss)[a,b] | 4 (0.7%) | 1 (0.2%) | 5 (0.5%) | 4 (0.7%) | 1 (0.2%) | 5 (0.5%) |
| Genito urinary system and sex hormones | 13 (2.3%) | 11 (2.3%) | 24 (2.3%) | 13 (2.3%) | 11 (2.3%) | 24 (2.3%) |
| Gynecologicals, sex hormones | 103 (18.1%) | 3 (0.6%) | 106 (10.0%) | 104 (18.2%) | 3 (0.6%) | 107 (10.1%) |
| Medication for treatment of chronic neurological diseases | 3 (0.5%) | 2 (0.4%) | 5 (0.5%) | 3 (0.5%) | 2 (0.4%) | 5 (0.5%) |
| Medications for allergy, allergens | 98 (17.2%) | 72 (14.8%) | 170 (16.1%) | 98 (17.1%) | 72 (14.8%) | 170 (16.1%) |
| Medications for asthma, cough and cold[c] | 7 (1.2%) | 4 (0.8%) | 11 (1.0%) | 7 (1.2%) | 4 (0.8%) | 11 (1.0%) |
| Medications for nervous system disorders (Anxiety, Depression, Bipolar) | 103 (18.1%) | 44 (9.1%) | 147 (13.9%) | 105 (18.4%) | 44 (9.1%) | 149 (14.1%) |
| Medications for treatment of arthritis and gout[d] | 0 (0.0%) | 6 (1.2%) | 6 (0.6%) | 0 (0.0%) | 6 (1.2%) | 6 (0.6%) |
| Medications for treatment of cardiovacular system (varicose veins) | 1 (0.2%) | 0 (0.0%) | 1 (0.1%) | 1 (0.2%) | 0 (0.0%) | 1 (0.1%) |
| Ophthalmologicals | 7 (1.2%) | 1 (0.2%) | 8 (0.8%) | 7 (1.2%) | 1 (0.2%) | 8 (0.8%) |
| Statins (prophylactic)[e] | 1 (0.2%) | 0 (0.0%) | 1 (0.1%) | 1 (0.2%) | 0 (0.0%) | 1 (0.1%) |
| Treatment of hypothyroidism or hyperthyoridism | 33 (5.8%) | 6 (1.2%) | 39 (3.7%) | 33 (5.8%) | 6 (1.2%) | 39 (3.7%) |

[a]Based on medical history (including body mass index), subjects were deemed eligible for participation
[b]One subject was prescribed Metformin for polyendocrine metabolic ovarian syndrome
[c]One subject with asthma was included but the study site PI deemed that the condition was not a major illness or chronic disease
[d]Subjects with gout were not deemed serious by the site PI and thus they were eligible for participation
[e]Statin taken based on heredity of high blood lipids; no medical history of high blood pressure prior to or during prescribed statin use

TABLE S4

**Table S4.** Overview of error messages observed in the 99th percentile and percent-over-LoD studies.

| **Error description** | **Error type** | **Events** (Total runs = 3225) | **% Total** |
|---|---|---|---|
| Sample volume low | User | 34 | 1.05 |
| Sample volume high | User | 37 | 1.15 |
| Cartridge use | User | 1 | 0.03 |
| Cartridge loading | User | 17 | 0.53 |
| Analyzer | Non-user | 2 | 0.06 |
| Cartridge processing | Non-user | 21 | 0.65 |
| Barcode reading | Non-user | 1 | 0.03 |
| | Total errors | 113 | 3.50% |

**Abbreviations:** LoD, limit of detection

TABLE S5

**Table S5.** SPINCHIP hs-cTnI limit of blank and limit of detection[a]

| Matrix | LoB ( ng/L) | LoD ( ng/L) |
|---|---|---|
| Plasma | 0.3 | 0.8 |
| WB | NA | 0.9 |

**Abbreviations:** hs-cTnI, high-sensitivity cardiac troponin I; LoB, limit of blank; LoD, limit of detection; WB, whole blood

[a]cTnI-depleted plasma was used as blank material, while low-level samples (≈1–5×LoB) were prepared from native and pooled plasma and fresh whole blood from healthy donors, with plasma samples frozen and whole blood analyzed on the day of collection

TABLE S6

**Table S6.** SPINCHIP hs-cTnI limit of quantitation (LoQ)[a]

| Matrix | LoQ; CV ≤20% ( ng/L) | LoQ; CV ≤10% ( ng/L) |
|---|---|---|
| Plasma | 1.1 | 3.8 |
| WB | 1.4 | 3.8 |

**Abbreviations:** hs-cTnI, high-sensitivity cardiac troponin I; CV, coefficient of variance; WB, whole blood

[a]Plasma and whole blood samples from healthy donors were screened for native cTnI, with selected samples spiked using a patient plasma pool to generate low-level concentration series; plasma was aliquoted and frozen, while whole blood was prepared with hematocrit-adjusted spiking and analyzed fresh on the day of collection

TABLE S7

**Table S7.** SPINCHIP hs-cTnI linearity[a]

| | Cartridge batch 1 | | | Cartridge batch 2 | | | Cartridge batch 3 | | |
|---|---|---|---|---|---|---|---|---|---|
| **Panel**[b] | **n** | **Target ( ng/L)** | **Bias** | **n** | **Target ( ng/L)** | **Bias** | **n** | **Target ( ng/L)** | **Bias** |
| 1 | 14 | 0.8 | 0.2 ng/L | 14 | 0.8 | 0.4 ng/L | 14 | 0.7 | 0.3 ng/L |
| 2 | 14 | 1.2 | 0.2 ng/L | 14 | 1.2 | 0.0 ng/L | 14 | 1.0 | 0.1 ng/L |
| 3 | 14 | 1.8 | 0.2 ng/L | 14 | 1.7 | 0.1 ng/L | 14 | 1.6 | 0.2 ng/L |
| 4 | 7 | 2.9 | 0.2 ng/L | 7 | 2.9 | 0.1 ng/L | 14 | 2.6 | 0.1 ng/L |
| 5 | 7 | 11.7 | -0.5 ng/L | 7 | 11.6 | -0.2 ng/L | 7 | 10.5 | 0.0 ng/L |
| 6 | 7 | 118 | 6% | 7 | 117 | 1% | 7 | 105 | 3% |
| 7 | 7 | 1152 | 4% | 7 | 1139 | 6% | 7 | 1031 | 2% |
| 8 | 7 | 3506 | 4% | 7 | 3468 | 3% | 7 | 3137 | 5% |
| 9 | 7 | 5657 | 5% | 7 | 5596 | 7% | 7 | 5062 | -2% |
| 10 | 7 | 767 | -8% | 7 | 7486 | -6% | 14 | 6772 | -7% |
| 11 | 7 | 10364 | -7% | 7 | 10252 | -7% | 7 | 9275 | -4% |

**Abbreviations:** hs-cTnI, high-sensitivity cardiac troponin I

[a]SPINCHIP hs-cTnI was considered linear if the deviation from linearity was ≤ ±3 ng/L or ≤ ±10%, whichever was the larger maximum

[b]Serial dilutions were prepared from a pooled high-troponin plasma sample (>6000 ng/L) using a troponin-free calibrator matrix as diluent, generating an 11-level panel spanning <1.3 to >9000 ng/L

**TABLE S8**

**Table S8.** Sample stability[a]

| | Sample ID | cTnI Baseline ( ng/L) | Difference (%) |
|---|---|---|---|
| ≥3 hours at 15-25ºC | Plasma 1 | 1250 | -3 |
| | Plasma 2 | 234 | -6 |
| | Plasma 3 | 1577 | -5 |
| | Plasma 4 | 29.3 | -5 |
| | Plasma 5 | 1181 | -1 |
| | Whole blood 1 | 263 | -4 |
| | Whole blood 2 | 65.8 | -6 |
| | Whole blood 3 | 2390 | -2 |
| | Whole blood 4 | 2426 | 2 |
| | Whole blood 5 | 5124 | 0 |
| ≥8 hours at 2-8ºC | Plasma 1 | 1250 | -6 |
| | Plasma 2 | 234 | -4 |
| | Plasma 3 | 1577 | -3 |
| | Plasma 4 | 29.3 | -9 |
| | Plasma 5 | 1181 | -6 |
| | Whole blood 1 | 263 | -7 |
| | Whole blood 2 | 65.8 | -4 |
| | Whole blood 3 | 2390 | 5 |
| | Whole blood 4 | 2426 | 3 |
| | Whole blood 5 | 5124 | 2 |
| ≥8 hours at -20ºC | Plasma 1 | 1107 | 0 |
| | Plasma 2 | 219 | -2 |
| | Plasma 3 | 1398 | -4 |
| | Plasma 4 | 26.4 | -1 |
| | Plasma 5 | 1027 | 5 |

**Abbreviations:** hs-cTnI, high-sensitivity cardiac troponin I;

[a]Fresh native whole blood and plasma samples, obtained from consenting patients, were used without spiking and analyzed over time under defined storage conditions to assess stability.

TABLE S9

**Table S9.** hs-cTnI shelf-life stability[a]

| | | | Change at $T_N$ across batches | | | Change at $T_{N+1}$ across batches | | |
|---|---|---|---|---|---|---|---|---|
| **Storage condition** | **Sample ID[b]** | **Shelf-life claim ($T_N$)** | **min** | **max** | ***10% beyond claim ($T_{N+1}$)*** | **min** | **max** | **Pass/Fail[c,d,e]** |
| 26 °C | Depleted (<1.1 ng/L) | 3 months | 0.0 ng/L | 0.1 ng/L | 3.4 months | 0.0 ng/L | 0.1 ng/L | Pass |
| | 5-15 ng/L | | 0.0 ng/L | 0.1 ng/L | | 0.0 ng/L | 0.1 ng/L | Pass |
| | 20-30 ng/L | | -1% | -2% | | -1% | -2% | Pass |
| | 500-1000 ng/L | | -3% | -6% | | -3% | -7% | Pass |
| 5 °C | Depleted (<1.1 ng/L) | 12 months | 0.0 ng/L | 0.0 ng/L | 13.2 months | 0.0 ng/L | 0.0 g/L | Pass |
| | 5-15 ng/L | | 0.0 ng/L | 0.3 ng/L | | 0.0 ng/L | -0.4 ng/L | Pass |
| | 20-30 ng/L | | -1% | -3% | | -1% | -3% | Pass |
| | 500-1000 ng/L | | -1% | -7% | | -1% | -8% | Pass |
| | *2-5 ng/L*[f] | | -0.1 ng/L | 0.1 ng/L | | -0.1 ng/L | 0.1 ng/L | Pass |
| | *100-300 ng/L*[f] | | -4% | -4% | | -5% | -5% | Pass |
| | *5000-9000 ng/L*[f] | | -3% | -6% | | -4% | -6% | Pass |

[a]Studies at longer timepoints are still ongoing for some batches; data will be updated prior to publication

[b]Plasma samples (depleted, native, and spiked to defined cTnI levels using patient plasma pools or recombinant TnITC) were prepared, aliquoted, and stored frozen until analysis for cartridge stability testing.

[c]Pass if drift from baseline ≤2 ng/L or ≤ 10 %, whichever is greater, for samples with Troponin I levels ≤ 3000 ng/L

[d]Pass if drift from baseline ≤15 % for samples with Troponin I levels >3000 ng/L

[e]Pass if Background signal (raw cTnI) <LoQ or increase from baseline ≤1 ng/L, for cTnI depleted samples

[f]Predicted based on regression analyses of interim results from batches 102103 and 102285.

TABLE S10

**Table S10.** Transport simulation stress sequence

| Stress test[a] | Details |
|---|---|
| Manual handling | Physical stress: 6 drops from 610 mm (Assurance level 1) |
| Cold climate (Freeze/thaw) | Environmental stress: 3 cycles of alternating storage at -18 ± 2 °C and 5 ± 3°C for ≥ 24 hours per temperature |
| Vibration test | Physical stress: Simulating transport by truck (60 min) and plane (120 min) up to 200 Hz |
| Low pressure (high altitude) | Physical stress: 1 hour at 50 ± 3 kPa, simulating transport by unpressurised airplane. |
| Manual handling | Physical stress: 6 drops from 610 mm |
| Tropical climate (40°C, 90% RH) | Environmental stress: Exposure to 40 ± 2 °C and 90 ± 5% RH for 3 days |

**Abbreviations:** RH, relative humidity

[a]Two batches were used for these experiments; one was exposed to the entire sequence, while the other was only exposed to the environmental stress tests

TABLE S11

**Table S11.** hs-cTnI transport simulation[a]

| | Stressed cartridges stored at 26 °C | | Stressed cartridges stored at 5 °C | |
|---|---|---|---|---|
| **Cartridge batch** | **Time from baseline** | **Pass/fail[a]** (across samples) | **Time from baseline** | **Pass/fail[a]** (across samples) |
| 101469 | 1 month | Pass | 3 months | Pass |
| 101469 | 2 months | Pass | 7 months | Pass |
| 101469 | 3 months | Pass | 9 months | Pass |
| 101469 | 3.4 months | Pass | 12 months | Pass |
| 101469 | 5 months | Pass | 13.2 months | Pass |
| 102420 | 1 month | Ongoing[c] | 3 months | Pass |
| 102420 | 3 months | Ongoing[c] | 5 months | Pass |
| 102420 | 3.4 months | Ongoing[c] | 9 months | Pass |

**Abbreviations:** hs-cTnI, high-sensitivity cardiac troponin I;

[a]Plasma samples were prepared from donor plasma, including native, spiked (patient pool or recombinant protein), and troponin-depleted matrices, then stored frozen and used for stability testing

[b]Pass if Background signal (raw cTnI) < LoQ or ≤ 1 ng/L, for cTnI depleted samples ; Pass if shift from unstressed ≤ 1 ng/L or ≤ 5 %, whichever is greater, for samples with Troponin I levels ≤ 3000 ng/L; Pass if shift from unstressed ≤ 7.5 % for samples with Troponin I levels > 3000 ng/L.

[c]Testing at later time points ongoing at the time of manuscript preparation.

**TABLE S12**

**Table S12.** Batch-to-batch variation for SPINCHIP hs-cTnI measurements

| | Mean overall bias | | |
|---|---|---|---|
| **cTnI interval[a]** | **Batch 01 vs. Batch 02** | **Batch 01 vs. Batch 03** | **Batch 02 vs. Batch 03** |
| 3 - 20 ng/L | 1 ng/L | 0 ng/L | -1 ng/L |
| 20 - 100 ng/L | 8% | 4% | -5% |
| 20 - 5000 ng/L | 8% | 6% | -2% |
| 20 - 9000 ng/L | 8% | 6% | -2% |
| **Slope** | 1.08 | 1.07 | 1.00 |
| **Intercept** | 0.00 | -0.98 | -0.85 |

**Abbreviations:** hs-cTnI, high-sensitivity cardiac troponin I;

[a]Samples for analysis were prepared primarily as native plasma samples (banked patient samples), with a small proportion of modified samples including plasma pools or single-donor plasma spiked with patient plasma

TABLE S13

**Table S13.** Potential Hook effect for recombinant cardiac troponin ITC complex and high levels of cTnI[a]

| Sample | Theoretical concentration ( ng/L) | Reported concentration ( ng/L) |
|---|---|---|
| Recombinant ITC samples | 1 000 000 | ≥ 9000 |
| | 100 000 | ≥ 9000 |
| | 10 000 | ≥ 9000 |
| **Sample** | **Architect cTnI concentration ( ng/L)** | **Reported concentration ( ng/L)** |
| High patient samples | 60 600 | ≥ 9000 |
| | 118 390 | ≥ 9000 |
| | 403 660 | ≥ 9000 |

**Abbreviations:** hs-cTnI, high-sensitivity cardiac troponin I

[a]Plasma samples were used, including (1) native single-donor plasma spiked with recombinant cardiac troponin ITC complex across a wide concentration range and (2) native patient plasma samples with very high endogenous cTnI concentrations.

**TABLE S14**

**Table S14.** Potential interference of SPINCHIP hs-cTnI performance due to the presence of human mouse antibodies (HAMA) or rheumatoid factor (RF) calculated as the relative difference in cTnI result between test samples and the control sample.

| | | HAMA (Activity)[a] | cTnI result [b]( ng/L) | Interference (%) |
|---|---|---|---|---|
| Control sample | | N/A | 41.4 | N/A |
| Test samples with HAMA | 1 | >5120 | 49.3 | 19 |
| | 2 | >1280 | 46.2 | 12 |
| | 3 | >5120 | 44.0 | 6 |
| | 4 | >1280 | 49.1 | 19 |
| | 5 | >2560 | 50.6 | 22 |
| | 6 | >400 | 41.8 | 1 |
| | 7 | >400 | 50.5 | 22 |
| | 8 | >400 | 49.1 | 19 |
| | 9 | >400 | 48.9 | 18 |
| | 10 | >400 | 45.1 | 9 |
| | | **RF (IU/mL)** | **cTnI result [b] ( ng/L)** | **Interference (%)** |
| Control sample | | N/A | 41.4 | N/A |
| Test samples with RF | 1 | 1820 | 37.2 | -10 |
| | 2 | 1710 | 46.7 | 13 |
| | 3 | 1770 | 46.1 | 11 |
| | 4 | 1740 | 44.4 | 7 |
| | 5 | 1710 | 42.8 | 4 |
| | 6 | 1770 | 44.5 | 8 |
| | 7 | 1740 | 42.7 | 3 |
| | 8 | 1660 | 50.4 | 22 |
| | 9 | 2941 | 47.1 | 14 |
| | 10 | 2330 | 47.6 | 15 |

**Abbreviations:** hs-cTnI, high-sensitivity cardiac troponin I

[a]Times more activity than a known negative

[b]cTnI concentration measured after spiking the sample (Test/Control) to a cTnI concentration between 30-100 ng/L (target 50 ng/L).

TABLE S15

**Table S15.** Potential interference of SPINCHIP hs-cTnI performance due to the presence of drugs[a]

| | Concentration | | | cTnI ( ng/L) | | | | |
|---|---|---|---|---|---|---|---|---|
| Drug | mg/dL | µmol/L | cTnI level | Control | n | Test | n | Interference (%) |
| Acetaminophen | | | Medium | 29.5 | 9 | 30.8 | 18 | 4 |
| | | | High | 699 | 18 | 687 | 9 | -2 |
| Acetylsalicylic acid | | | Medium | 29.2 | 18 | 29.5 | 9 | 1 |
| | | | High | 666 | 9 | 685 | 9 | 3 |
| Ampicillin | | | Medium | 38.4 | 18 | 38.4 | 9 | 0 |
| | | | High | 805 | 9 | 784 | 9 | -3 |
| Ascorbic Acid | | | Medium | 38.6 | 9 | 37.3 | 18 | -3 |
| | | | High | 807 | 9 | 790 | 9 | -2 |
| Atenolol | | | Medium | 29.6 | 18 | 30.4 | 18 | 3 |
| | | | High | 688 | 9 | 713 | 9 | 4 |
| Atorvastatin | | | Medium | 34.9 | 9 | 36.9 | 9 | 6 |
| | | | High | 745 | 18 | 766 | 9 | 3 |
| Biotin | | | Medium | 31.3 | 9 | 32.5 | 9 | 4 |
| | | | High | 673 | 9 | 664 | 9 | -1 |
| Caffeine | | | Medium | 38.6 | 9 | 40.4 | 9 | 5 |
| | | | High | 807 | 9 | 791 | 9 | -2 |
| Captopril | | | Medium | 38.4 | 18 | 39.0 | 18 | 2 |
| | | | High | 805 | 9 | 804 | 9 | 0 |
| Carvedilol | | | Medium | 34.9 | 9 | 35.7 | 9 | 2 |
| | | | High | 745 | 18 | 749 | 9 | 1 |
| Cefoxitin[b] | | | Medium | 33.6 | 9 | 25.6 | 18 | -24 |
| | | | High | 740 | 18 | 544 | 9 | -26 |
| Cefoxitin 100 % (Titration) | | | Medium | 34.0 | 18 | 25.1 | 18 | -26 |
| | | | High | 741 | 9 | 562 | 9 | -24 |
| Cefoxitin 75 % (Titration) | | | Medium | 34.0 | 18 | 26.9 | 18 | -21 |
| | | | High | 741 | 9 | 600 | 9 | -19 |
| Cefoxitin 75 % (Titration) | | | Medium | 34.0 | 18 | 28.6 | 18 | -16 |
| | | | High | 741 | 9 | 633 | 9 | -15 |
| Cefoxitin 25 % (Titration) | | | Medium | 34.0 | 18 | 30.7 | 17 | -10 |
| | | | High | 741 | 9 | 675 | 9 | -9 |
| Clopidogrel | | | Medium | 29.9 | 9 | 31.0 | 18 | 4 |
| | | | High | 696 | 9 | 711 | 9 | 2 |
| Codeine | | | Medium | 36.1 | 18 | 36.7 | 18 | 2 |
| | | | High | 757 | 9 | 787 | 9 | 4 |
| Diclofenac | | | Medium | 31.1 | 18 | 31.6 | 18 | 2 |
| | | | High | 714 | 18 | 704 | 9 | -1 |
| Digoxin | | | Medium | 34.0 | 9 | 33.1 | 18 | -3 |
| | | | High | 699 | 9 | 761 | 9 | 9 |
| Dopamine | | | Medium | 38.4 | 18 | 39.2 | 18 | 2 |
| | | | High | 805 | 9 | 809 | 9 | 0 |
| Doxycycline | | | Medium | 32.1 | 9 | 32.1 | 18 | 0 |
| | | | High | 708 | 18 | 710 | 18 | 0 |
| Ethanol | | | Medium | 38.4 | 9 | 37.0 | 18 | -4 |
| | | | High | 805 | 9 | 797 | 9 | -1 |
| Fibrinogen | | | Medium | 33.6 | 18 | 34.9 | 9 | 4 |
| | | | High | 741 | 18 | 801 | 9 | 8 |

Table S15 (continued)

| | Concentration | | | cTnI ( ng/L) | | | | |
|---|---|---|---|---|---|---|---|---|
| Drug | mg/dL | µmol/L | cTnI level | Control | n | Test | n | Interference (%) |
| Furosemide | | | Medium | 33.9 | 18 | 34.3 | 18 | 1 |
| | | | High | 695 | 9 | 698 | 9 | 0 |
| Ibuprofen | | | Medium | 29.7 | 9 | 30.1 | 9 | 1 |
| | | | High | 685 | 9 | 700 | 9 | 2 |
| Insulin | | | Medium | 37.2 | 9 | 38.0 | 9 | 2 |
| | | | High | 811 | 9 | 820 | 9 | 1 |
| Lisinopril | | | Medium | 38.6 | 9 | 39.1 | 9 | 1 |
| | | | High | 807 | 9 | 798 | 9 | -1 |
| L—Nicotine | | | Medium | 32.6 | 18 | 32.7 | 18 | 0 |
| | | | High | 735 | 9 | 737 | 9 | 0 |
| LMW heparin | | | Medium | 34.9 | 9 | 34.2 | 9 | -2 |
| | | | High | 745 | 18 | 737 | 9 | -1 |
| Methyldopa | | | Medium | 38.6 | 9 | 37.4 | 9 | -3 |
| | | | High | 807 | 9 | 813 | 9 | 1 |
| Metformin | | | Medium | 33.2 | 9 | 32.9 | 9 | -1 |
| | | | High | 729 | 18 | 733 | 9 | 1 |
| Nitrofurantoin | | | Medium | 29.3 | 9 | 29.2 | 18 | 0 |
| | | | High | 669 | 9 | 695 | 9 | 4 |
| Propranolol | | | Medium | 30.0 | 18 | 30.4 | 9 | 1 |
| | | | High | 664 | 9 | 653 | 9 | -2 |
| Rosuvastatin | | | Medium | 34.9 | 9 | 35.0 | 9 | 0 |
| | | | High | 745 | 18 | 760 | 9 | 3 |
| Simvastatin | | | Medium | 29.6 | 9 | 29.2 | 18 | -2 |
| | | | High | 631 | 9 | 614 | 9 | -3 |
| Streptokinase | | | Medium | 32.9 | 18 | 32.6 | 9 | -1 |
| | | | High | 682 | 9 | 666 | 18 | -2 |
| Theophylline | | | Medium | 37.0 | 9 | 36.5 | 18 | -1 |
| | | | High | 798 | 9 | 797 | 9 | 0 |
| Valsartan | | | Medium | 34.9 | 9 | 33.8 | 9 | -3 |
| | | | High | 745 | 18 | 762 | 9 | 2 |
| Verapamil | | | Medium | 30.2 | 9 | 31.5 | 18 | 4 |
| | | | High | 651 | 9 | 658 | 9 | 1 |
| Warfarin | | | Medium | 31.6 | 9 | 32.1 | 9 | 1 |
| | | | High | 695 | 9 | 667 | 9 | -4 |

**Abbreviations:** hs-cTnI, high-sensitivity cardiac troponin I

[a]Pooled plasma derived from banked patient samples was used, specifically medium- and high-level plasma pools (≈15–60 ng/L and 500–1500 ng/L cTnI), which were spiked with drugs (and matched solvent controls).
[b]No interference is defined as a result ≤ ±10% or a bias ≤ ± 2 ng/L relative to a control sample

TABLE S16

**Table S16.** Potential interference of SPINCHIP hs-cTnI performance due to the presence of bilirubin and hemoglobin[a]

| Endogenous substance | Concentration | cTnI level | cTnI ( ng/L) | | | | Interference (%) |
|---|---|---|---|---|---|---|---|
| | | | Control | n | Test | n | |
| Conjugated bilirubin | 475 µmol/L | Medium | 35.6 | 9 | 33.4 | 18 | -6 |
| | | High | 705 | 9 | 675 | 9 | -4 |
| Unconjugated bilirubin | 684 µmol/L | Medium | 32.2 | 18 | 29.9 | 18 | -7 |
| | | High | 675 | 9 | 626 | 9 | -7 |
| Hemoglobin | 10.21 g/L | Medium | 33.4 | 9 | 34.9 | 9 | 5 |
| | | High | 652 | 9 | 691 | 9 | 6 |

TABLE S17

**Table S17.** Potential endogenous interference of SPINCHIP hs-cTnI performance due to the presence of thrombocytes[a]

| | | cTnI ( ng/L) | | |
|---|---|---|---|---|
| **Sample ID** | **Number of thrombocytes (platelets/L)[b]** | **Control plasma** | **Whole blood** | **Interference** |
| 01 | $315 \times 10^9$ | 910 | 933 | 3 % |
| 02 | $335 \times 10^9$ | 7.9 | 8.2 | 6 % |
| 03 | $370 \times 10^9$ | 1121 | 1114 | -1 % |
| 04 | $370 \times 10^9$ | 2.2 | 3.0 | 0.7 ng/L |
| 05 | $415 \times 10^9$ | 633 | 667 | 5 % |
| 06 | $428 \times 10^9$ | 31.9 | 29.3 | -8 % |
| 07 | $441 \times 10^9$ | 931 | 1018 | 9 % |
| 08 | $465 \times 10^9$ | 6.3 | 6.6 | 0.3 ng/L |
| 09 | $490 \times 10^9$ | 2.7 | 2.7 | 0.0 ng/L |
| 10 | $491 \times 10^9$ | 766 | 771 | 1 % |

**Abbreviations:** hs-cTnI, high-sensitivity cardiac troponin I

[a]Whole blood patient samples with elevated thrombocyte (platelet) levels (≈300–500 $\times 10^9$/L) were used, along with matched control plasma prepared from the same samples after removal of thrombocytes by centrifugation. For cTnI, samples were either spiked with recombinant Troponin ITC (500–1500 ng/L) or used with native cTnI (>2 ng/L), with paired thrombocyte-depleted plasma controls prepared by centrifugation
[b]Control plasma was prepared from each whole blood sample by centrifugation at 2500 x g for 15 min to remove most of the thrombocytes present.

**TABLE S18**

**Table S18.** Potential endogenous interference of SPINCHIP hs-cTnI performance due to the presence of hematocrit[a]

| Sample type | | Hct level (%) | cTnI ( ng/L) | Interference (%) | Relevant Error messages (n) |
|---|---|---|---|---|---|
| Whole blood Donor 1 | Plasma control sample | | 116 | | |
| | Hct test sample level 1 | 52 | 108 | -7 | 0 |
| | Hct test sample level 2 | 53 | 112 | -3 | 0 |
| | Hct test sample level 3 | 55 | 111 | -4 | 0 |
| | Hct test sample level 4 | 58 | 113 | -3 | 2 |
| Whole blood Donor 2 | Plasma control sample | | 153 | | |
| | Hct test sample level 1 | 46 | 144 | -6 | 0 |
| | Hct test sample level 2 | 50 | 144 | -6 | 0 |
| | Hct test sample level 3 | 53 | 144 | -6 | 0 |
| | Hct test sample level 4 | 55 | 143 | -7 | 0 |
| Whole blood Donor 3 | Plasma control sample | | 120 | | |
| | Hct test sample level 1 | 43 | 119 | -1 | 0 |
| | Hct test sample level 2 | 62 | 111 | -7 | 3 |
| Whole blood Donor 4 | Plasma control sample | | 130 | | |
| | Hct test sample level 1 | 45 | 130 | 0 | 0 |
| | Hct test sample level 2 | 64 | 129 | -1 | 2 |

**Abbreviations:** Hct, hematocrit; hs-cTnI, high-sensitivity cardiac troponin I

[a]Whole blood samples spiked with recombinant cardiac troponin ITC were aliquoted, with plasma removed after sedimentation to generate a range of hematocrit levels (~46–64%), alongside matched plasma controls

**TABLE S19**

**Table S19.** Potential endogenous interference of SPINCHIP hs-cTnI performance due to the presence of lipids[a]

| | | | cTnI ( ng/L) | | |
|---|---|---|---|---|---|
| **Substance** | **Concentration** | **cTnI level** | **Control** | **Test** | **Interference (%)** |
| Intralipid[b] | 2000 mg/dL | Medium | 30.8 | 30.1 | -2 |
| | | High | 627 | 592 | -6 |

**Abbreviations:** hs-cTnI, high-sensitivity cardiac troponin I

[a]Plasma samples prepared from pooled patient specimens were used at two cardiac troponin I levels (~30 ng/L and ~600 ng/L), with aliquots spiked with Intralipid (test) or saline (control) to evaluate lipid interference
[b]Intralipid samples were prepared from an emulsion of soybean oil, which consists of a mixture of neutral triglycerides and predominantly unsaturated fatty acids

TABLE S20

**Table S20.** Potential endogenous interference of SPINCHIP hs-cTnI performance due to the presence of leukocytes[a]

| | | cTnI ( ng/L) | | |
|---|---|---|---|---|
| **Sample ID** | **Number of leukocytes (cells/L)[b]** | **Control plasma** | **Whole blood** | **Absolute difference ( ng/L)** |
| 01 | $10.5 \times 10^9$ | 6.5 | 6.9 | 0.4 |
| 02 | $12.5 \times 10^9$ | 5.7 | 7.0 | 1.3 |
| 03 | $14.0 \times 10^9$ | 15.4 | 15.7 | 0.3 |
| 04 | $15.3 \times 10^9$ | 8.4 | 8.9 | 0.5 |
| 05 | $17.9 \times 10^9$ | 8.5 | 8.4 | -0.1 |
| 06 | $19.2 \times 10^9$ | 12.5 | 12.7 | 0.2 |

**Abbreviations:** hs-cTnI, high-sensitivity cardiac troponin I

[a]Whole blood patient samples with elevated leukocyte levels (10.5–19.2 × $10^9$ cells/L) and native cardiac troponin I concentrations (~5–15 ng/L) were analyzed alongside matched leukocyte-reduced plasma prepared from the same specimens to assess leukocyte interference
[b]Control plasma was prepared from a fraction of each whole blood sample through centrifugation at 2500 x g for 15 minutes, to remove most of the leukocytes present. ng/L

**TABLE S21**

**Table S21.** Potential cross-reactivity of skTnI

| | | Cartridge batch | Cross-reactivity (%) |
|---|---|---|---|
| 1 000 000 ng/L skTnI[a] | Depleted plasma (<LoQ) | 100973 | 0.021 |
| | | 100999 | 0.021 |
| | | 101028 | 0.029 |
| | Medium base pool (15-60 ng/L) | 100973 | 0.021 |
| | | 100999 | 0.020 |
| | | 101028 | 0.029 |
| | High base pool (500-1500 ng/L) | 100973 | 0.023 |
| | | 100999 | 0.027 |
| | | 101028 | 0.038 |

**Abbreviations:** skTnI, skeletal troponin I; LoQ, limit of quantification

[a]Samples prepared from native plasma samples (healthy donor and patient pools) that were spiked with recombinant skeletal troponin I (skTnI)

TABLE S22

**Table S22.** Potential cross-reactivity of cTnC and cTnT

| Test concentration | Sample[a] | Cartridge batch | Cross-reactivity (%) |
|---|---|---|---|
| 1 000 000 ng/L cTnC | Depleted plasma (<1.1 ng/L) | 100973 | < 0.001 |
| | | 100999 | < 0.001 |
| | | 101028 | < 0.001 |
| | Medium base pool (15-60 ng/L) | 100973 | 0.001 |
| | | 100999 | 0.001 |
| | | 101028 | 0.001 |
| | High base pool (500-1500 ng/L) | 100973 | 0.016 |
| | | 100999 | 0.015 |
| | | 101028 | 0.012 |
| 1 000 000 ng/L cTnT | Depleted plasma (<1.1 ng/L) | 100973 | < 0.001 |
| | | 100999 | < 0.001 |
| | | 101028 | < 0.001 |
| | Medium base pool (15-60 ng/L) | 100973 | < 0.001 |
| | | 100999 | < 0.001 |
| | | 101028 | < 0.001 |
| | High base pool (500-1500 ng/L) | 100973 | < 0.001 |
| | | 100999 | < 0.001 |
| | | 101028 | < 0.001 |

**Abbreviations:** cTnC, cardiac troponin C; cTnT, cardiac troponin T

[a]Native plasma samples (a cTnI-depleted pool and patient plasma pools at medium and high cTnI levels) were spiked with cardiac troponin C or T to a final concentration of $1X10^{6}$ ng/L, while matched control samples were prepared by adding an equal volume of troponin-free control solution

**TABLE S23**

**Table S23.** Potential cross-reactivity of creatine kinase myocardial band

| Test concentration | Sample[a] | Cartridge batch | Cross-reactivity (%) |
|---|---|---|---|
| 1 000 000 ng/L CK-MB | Depleted plasma (<1.1 ng/L) | 100973 | 0.002 |
| | | 100999 | 0.002 |
| | | 101028 | 0.002 |
| | Medium base pool (15-60 ng/L) | 100973 | 0.002 |
| | | 100999 | 0.002 |
| | | 101028 | 0.002 |
| | High base pool (500-1500 ng/L) | 100973 | 0.002 |
| | | 100999 | 0.003 |
| | | 101028 | 0.003 |

**Abbreviations:** CK-MB, creatine kinase myocardial band

[a]Native plasma samples (a cTnI-depleted sample and patient plasma pools at medium and high cTnI levels) were prepared by spiking them with CK-MB to a final concentration of $1X10^{6}$ ng/L, with matched control samples receiving an equal volume of CK-MB-free control solution

**TABLE S24**

**Table S24.** Potential cross-reactivity of myoglobin

| Concentration | Sample[a] | Cartridge batch | Cross-reactivity (%) |
|---|---|---|---|
| 1 000 000 ng/L Myoglobin | Depleted plasma (<1.1 ng/L) | 100973 | < 0.001 |
| | | 100999 | < 0.001 |
| | | 101161 | < 0.001 |
| | Medium base pool (15-60 ng/L) | 100973 | < 0.001 |
| | | 100999 | < 0.001 |
| | | 101161 | < 0.001 |
| | High base pool (500-1500 ng/L) | 100973 | 0.001 |
| | | 100999 | 0.002 |
| | | 101161 | 0.002 |

[a]Native plasma samples (a cTnI-depleted sample and patient plasma pools at medium and high cTnI levels) were prepared by spiking them with myoglobin to a final concentration of $1X10^6$ ng/L, with matched control samples receiving an equal volume of myoglobin-free control solution

TABLE S25

**Table S25.** SPINCHIP hs-cTnI repeatability (within-run) precision for plasma and whole blood samples; and within-laboratory precision for plasma samples[a]

| | | Plasma | | | | | | Whole blood | | | |
|---|---|---|---|---|---|---|---|---|---|---|---|
| | | | | Repeatability | | Within-laboratory | | | | Repeatability[b] | |
| Sample | Cartridge | n | Mean | SD (ng/L) | CV (%) | SD (ng/L) | CV (%) | n | Mean | SD (ng/L) | CV (%) |
| 1 | a | 80 | 10.5 | 0.7 | 6.8 | 0.7 | 6.8 | 40 | 14.4 | 1.0 | 7.1 |
| | b | 80 | 9.7 | 0.7 | 7.5 | 0.8 | 7.7 | 40 | 9.1 | 0.6 | 6.5 |
| | c | 80 | 9.2 | 0.5 | 5.8 | 0.6 | 6.5 | 40 | 9.3 | 0.7 | 8.0 |
| 2 | a | 80 | 30.0 | 1.5 | 4.9 | 1.5 | 5.1 | 40 | 30.9 | 1.6 | 5.3 |
| | b | 80 | 27.1 | 1.0 | 3.8 | 1.1 | 4.1 | 40 | 25.4 | 1.3 | 5.3 |
| | c | 80 | 25.3 | 1.0 | 4.1 | 1.4 | 5.6 | 40 | 24.8 | 1.1 | 4.3 |
| 3 | a | 80 | 170 | 7.5 | 4.4 | 7.8 | 4.6 | 40 | 173 | 7.9 | 4.6 |
| | b | 80 | 149 | 5.6 | 3.8 | 6.2 | 4.2 | 40 | 165 | 5.3 | 3.2 |
| | c | 80 | 133 | 4.3 | 3.2 | 5.0 | 3.7 | 40 | 147 | 5.6 | 3.8 |
| 4 | a | 80 | 1968 | 89.4 | 4.5 | 94.1 | 4.8 | 40 | 1588 | 90 | 5.6 |
| | b | 80 | 1806 | 71.7 | 4.0 | 87.9 | 4.9 | 40 | 1571 | 83 | 5.3 |
| | c | 80 | 1939 | 81.4 | 4.2 | 85.9 | 4.4 | 40 | 1738 | 81 | 4.7 |

**Abbreviation:** WB, whole blood; SD, standard deviation; CV, coefficient of variation;

[a]Plasma and whole blood samples were prepared by spiking donor samples with patient plasma or recombinant troponin to defined concentration levels, with plasma samples aliquoted and stored frozen until analysis
[b]Whole blood samples were freshly prepared, and analyzed the same day due to stability constraints

TABLE S26

**Table S26.** Within-run precision (repeatability), between-day precision, between-instrument precision and reproducibility for specimens collected for a multi-instrument precision study[a]

| | | | Repeatability | | Between-day | | Between-instrument | | Reproducibility | |
|---|---|---|---|---|---|---|---|---|---|---|
| **Sample** | **n** | **Mean cTnI ( ng/L)** | **SD ( ng/L)** | **CV (%)** | **SD ( ng/L)** | **CV (%)** | **SD ( ng/L)** | **CV (%)** | **SD ( ng/L)** | **CV (%)** |
| 1 | 250 | 8.4 | 1 | 9 | 0 | 2 | 0 | 2 | 1 | 9 |
| 2 | 250 | 30.3 | 2 | 5 | 0 | 1 | 1 | 2 | 2 | 6 |
| 3 | 250 | 309 | 12 | 4 | 4 | 1 | 7 | 2 | 14 | 5 |
| 4 | 250 | 4481 | 206 | 5 | 60 | 1 | 131 | 3 | 251 | 6 |

**Abbreviations:** cTnI, cardiac troponin I; SD, standard deviation; CV, coefficient of variation

[a]Plasma samples were prepared from donor plasma, with three samples spiked using patient plasma pools or recombinant troponin to achieve target concentrations while one was used native.

TABLE S27

**Table S27.** Within-run precision (repeatability), within-laboratory precision, and between-site reproducibility for plasma specimens collected for a multi-site precision study[a]

| | | | Repeatability | | Within-laboratory | | Between-site | |
|---|---|---|---|---|---|---|---|---|
| **Sample** | **n** | **Mean cTnI ( ng/L)** | **SD ( ng/L)** | **CV (%)** | **SD ( ng/L)** | **CV (%)** | **SD ( ng/L)** | **CV (%)** |
| 1 | 75 | 3.7 | 0.6 | 16.5 | 0.6 | 17.2 | 0.63 | 17.2 |
| 2 | 75 | 19.8 | 1.6 | 8.2 | 1.6 | 8.2 | 1.70 | 8.5 |
| 3 | 75 | 235 | 15.7 | 6.6 | 15.75 | 6.6 | 19.46 | 8.3 |
| 4 | 75 | 2699 | 115 | 4.3 | 115 | 4.3 | 115.02 | 4.3 |
| 5 | 75 | 7657 | 331 | 4.3 | 338 | 4.4 | 390.04 | 5.1 |

**Abbreviations:** cTnI, cardiac troponin I; SD, standard deviation; CV, coefficient of variation

[a]Plasma samples from adult subjects with troponin I concentrations spanning five predefined levels (~5–9000 ng/L) were analyzed across multiple sites, alongside commercial control materials, to evaluate multisite precision of the assay

TABLE S28

**Table S28.** Within-day (repeatability) SPINCHIP hs-cTnI results for whole blood samples collected for a multi-site precision study[a]

| | | | | Repeatability | |
|---|---|---|---|---|---|
| **Sample** | **Site** | **n** | **Mean cTnI ( ng/L)** | **SD ( ng/L)** | **CV (%)** |
| 1 | 1 | 40 | 3.7 | 0.5 | 13.9 |
| 1 | 2 | 40 | 3.7 | 0.5 | 12.6 |
| 2 | 3 | 40 | 6.9 | 0.6 | 8.2 |
| 3 | 1 | 40 | 20.2 | 1.4 | 6.9 |
| 3 | 2 | 40 | 20.2 | 1.3 | 6.3 |
| 4 | 3 | 40 | 24.0 | 1.6 | 6.6 |
| 5 | 1 | 40 | 251 | 13.6 | 5.4 |
| 5 | 2 | 40 | 253 | 8.2 | 3.3 |
| 6 | 3 | 40 | 145 | 6.0 | 4.2 |
| 7 | 1 | 40 | 2692 | 91.7 | 3.4 |
| 7 | 2 | 40 | 2660 | 89.1 | 3.4 |
| 8 | 3 | 40 | 2291 | 95.3 | 4.2 |
| 9 | 1 | 40 | 7934 | 500 | 6.3 |
| 9 | 2 | 40 | 7787 | 297 | 3.8 |
| 10 | 3 | 40 | 5619 | 244 | 4.3 |

**Abbreviations:** hs-cTnI, high-sensitivity cardiac troponin I; SD, standard deviation; CV, coefficient of variation

[a]Whole blood samples from adult subjects with troponin I concentrations spanning five predefined levels (~5–9000 ng/L) were analyzed across multiple sites, alongside commercial control materials, to evaluate multisite precision of the assay

**SUPPLEMENTARY FIGURES**
**FIGURE S1**

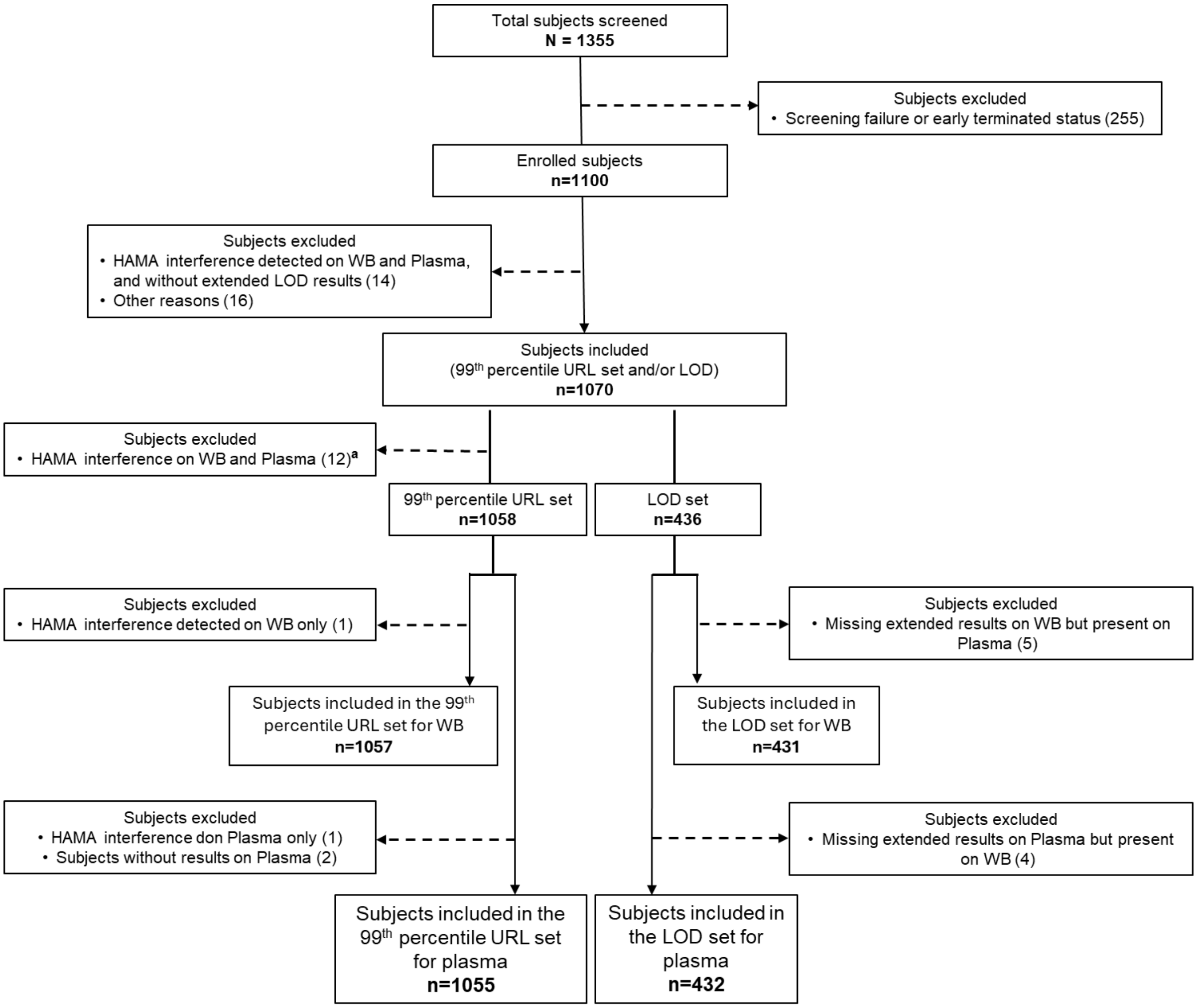


**Figure S1.** Specimen reconciliation for the per-protocol population used in the determination of the 99th percentile upper reference limit and in assessing the proportion of healthy subjects with troponin I concentrations above the limit of detection is presented. Reconciliation summaries are provided separately for plasma and whole blood samples.
[a]Represent the same samples

FIGURE S2

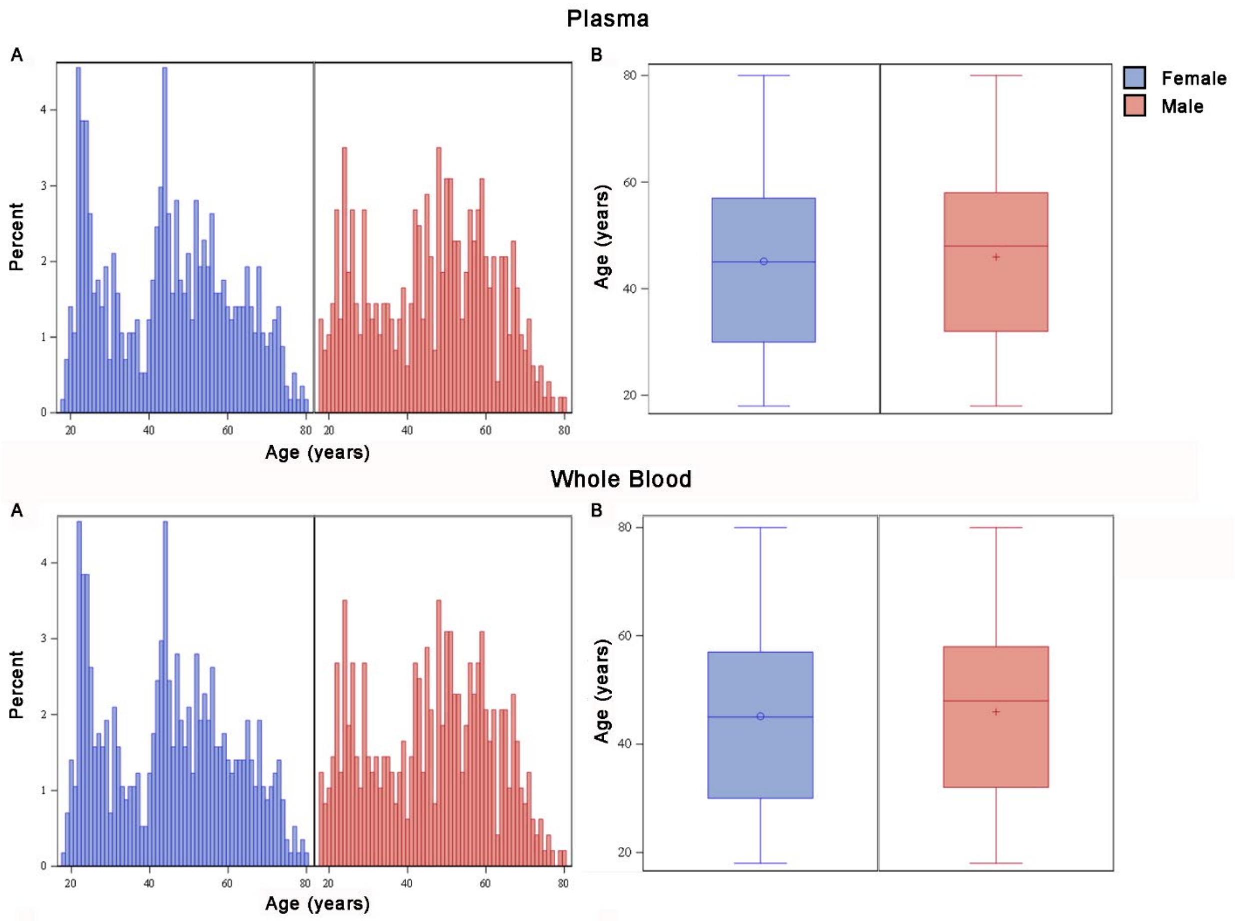


**Figure S2.** Age distribution in the 99th percentile population by sex. For plasma samples, N=1055; for whole blood, N=1057.

FIGURE S3

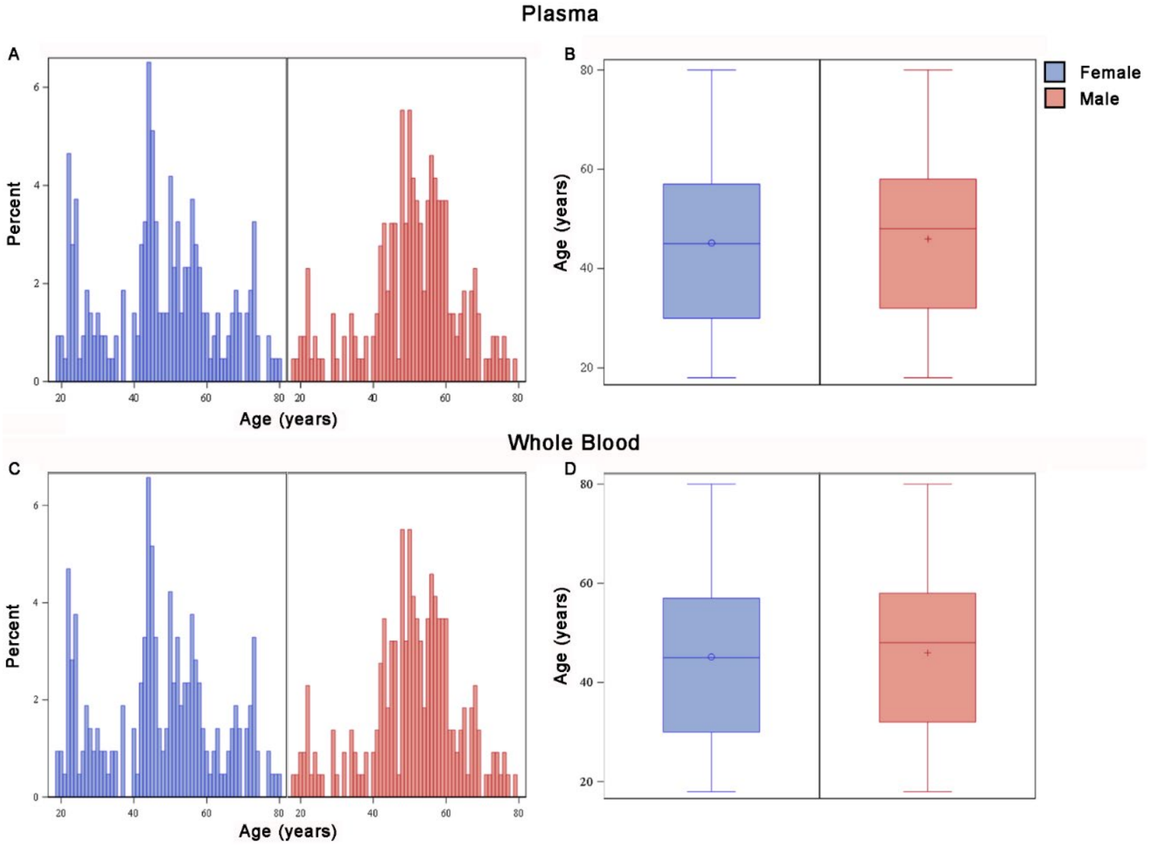


**Figure S3.** Age distribution in the LoD population by sex. For plasma samples, N=432; for whole blood, N=431.

**FIGURE S4**

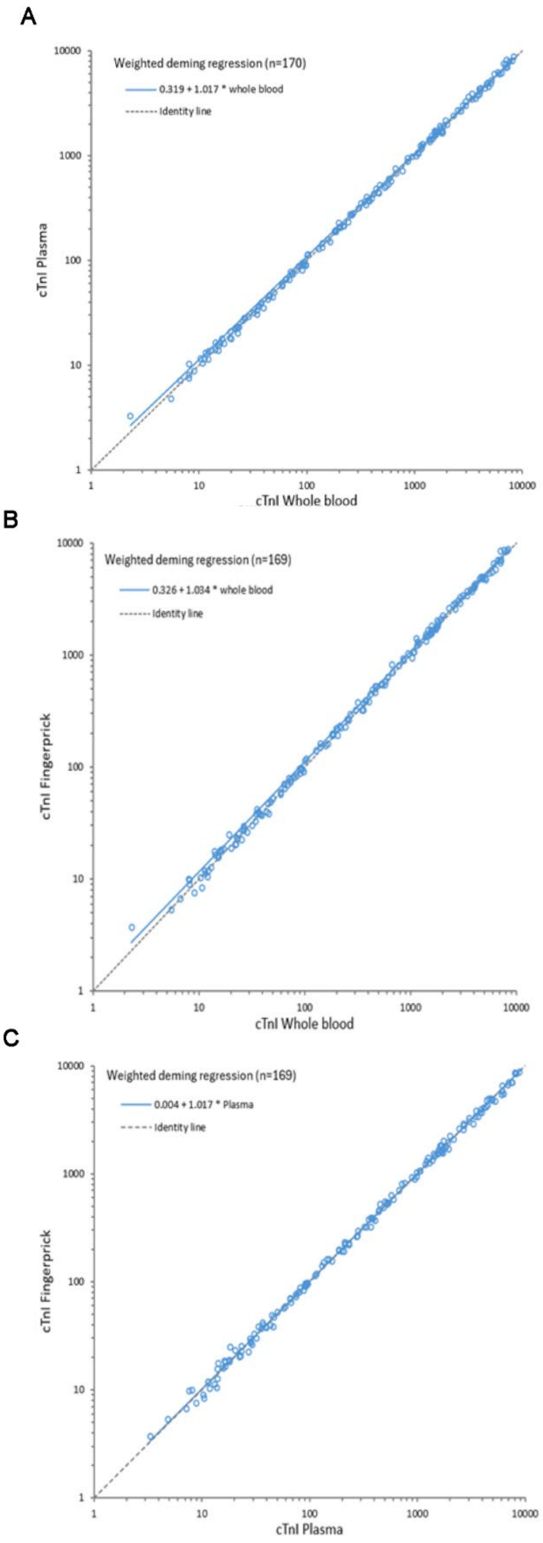


**FIGURE S4.** Weighted Deming regression (A) plasma versus whole blood; (B) finger-prick (capillary blood) versus whole blood; (C) finger prick versus plasma.
